# Measuring Planetary Eco-Emotions: A Systematic Review of Currently Available Instruments and Their Psychometric Properties

**DOI:** 10.1101/2024.03.22.24304713

**Authors:** Fulya Kırımer-Aydınlı, Mariel Juaréz Castelán, Nilab Hakim, Pelin Gul, A. Berfu Unal, Raimundo Aguayo-Estremera, Adriana Perez Fortis, Mario E. Rojas-Russell, Valentina Gallo

## Abstract

**Background:** The climate crisis has a wide range of direct and indirect mental health impacts on populations. However, their quantification is limited by the lack of unified definitions and assessment tools. The aim of this systematic review is to map all psychometric instruments used to measure emotions associated with the climate crisis, evaluate their psychometric characteristics, and identify any existing gaps.

**Methods:** The protocol was registered on PROSPERO. Data were reported following the COSMIN Risk of Bias of PROM and PRISMA checklists. Original articles describing the psychometric properties and/or validation of self-report measures designed to assess eco-anxiety and other climate change-related emotions in the general population were within the scope of this review. PubMed, PsycINFO, and Web of Science were the search engines used.

**Findings:** A total of 10 different psychometric scales measuring various eco-emotions were identified. Four focused on anxiety, while the remaining six focused on combinations of other negative emotions. The definitions of eco-emotions were not consistent across papers. Most of the instruments were developed in the Global North. Six of the instruments were multidimensional. All but one scale included at least one item indicating behavioural, cognitive, or physical aspects of emotions toward climate crises. The most recurrent emotion was worry, followed by anxiety, fear, and sadness. Including ten scale development studies, a total of 22 studies reporting instrument validation were reviewed. Two of the instruments have been validated in other populations.

**Interpretation:** To what extent the emotions covered by the instruments may overlap in relation to climate change is, to date, not clear. This is due to the lack of consistent definitions of climate-related emotions. Moreover, the mention of emotions was derived by a top-down approach, in all included studies. No positive emotions, such as hopefulness, humor, anticipated pride, gratitude, optimism, or feeling strong to do something though own contributions, have been detected.

## Introduction

Climate change refers to the ecological crisis caused by human-induced rising temperatures on the planet.^1^ It is widely recognized as one of the most serious global health threats of the 21^st^ century, menacing public health worldwide, affecting the physical and mental health of populations alike.^1,2^

The impacts of climate change on mental health can result from both direct and indirect exposure to extreme climate events. Direct exposure takes place when a person experiences first-hand extreme weather events such as bushfires or flooding, or food and water shortages because of climate change. Indirect impacts include short- and long-term losses in the economy (e.g., decrease in the yield of crops grown due to changes in the ecosystem) as well as physiological (e.g., infectious diseases) and psychological (e.g., the sense of anxiety leading to psychological trauma) consequences caused by extreme weather events.^3–5^ Studies show how both direct and indirect exposure to extreme climate events have a negative impact on populations’ mental health.^6,7^ In recent years, the number of studies on the mental health consequences of climate change has increased dramatically. A rapid search of PubMed conducted on the 6^th^ of February 2024 using the terms ‘climate change’ and ‘mental health’ yielded over 800 results, the majority of which published in the past five years reaching the maximum number in 2023. The focus has been mostly placed on understanding and quantifying which types of emotions are triggered by the environmental crisis.^8–12^

Some emotions explored so far include eco-anxiety, eco-guilt, eco-grief, solastalgia, and eco-depression ^13–17^ (Box 1). Eco-anxiety or climate change anxiety is one of the most frequently studied.^9,18–25^ Climate change anxiety, climate anxiety, climate change worry, or environmental anxiety are often used interchangeably; in some studies, however, it was suggested that climate change anxiety is a subset of eco-anxiety.^17^ Eco-anxiety and eco-grief exhibit distinct characteristics: eco-anxiety is primarily focused on the future, reacting to anticipated ecological losses, while eco-grief is more centred around real or past ecological losses, or reactions to future situations causing current losses^14^. Eco-grief resonates strongly with solastalgia.^26^ Often eco-anxiety has been used as an umbrella term capturing all these related emotions.^22,27^ Yet disagreements regarding the definition of this concept continue^14^ implying that the understanding of these phenomena is still in the process of development.^28^ Other concepts that encapsulate some eco-emotions are also found in the literature, such as eco-coping, eco-concern, and eco-uncertainty (Box 1).

Climate change’s impact on mental health is a growing but still underdeveloped area of research. It is imperative to accelerate and broaden research efforts to formulate evidence-based strategies for mitigation and adaptation.^29^ This can be achieved by revealing all the emotions that motivate people to take action.^30^ Consequently, very recently, the focus has been shifted from eco-anxiety or eco-anger to studying eco-emotions, as a way of conceptually characterizing the various types of emotions.^30^ Emotions are also tightly associated with cognitive and behaviour traits, which are sometime used to assess the emotions themselves. For example, one could ask to what extent a person perceives eco-anxiety (the emotion), or if they have negative perspectives on the future of the planet because of climate change (cognition), or if they sleep poorly because of climate change (behaviour).^15,17^

The aim of this systematic review is to map all instruments used to measure eco-emotions associated with the climate crisis, appraise their psychometric characteristics, and identify any existing gap. In doings so, this systematic review also contributes to advocating for the need of more consistent definitions of eco-emotions in the future.

## Method

### Protocol design and Registration

A systematic review protocol was developed and registered on PROSPERO (ID Number: CRD42023421638) on the 25^th^ of May 2023. The updated protocol adapted to the content of the literature retrieved is reported below. No ethical approval was sought for this study as it includes only publicly available data. Data are reported following the COSMIN-RoB (Risk of Bias of PROM validity studies^31^) and PRISMA checklist.^32^

### Source of Data

The search terms were developed using the broad fields of eco-emotions and psychometric instruments linked with Boolean connectors. A librarian from the University of Groningen was consulted for refining the search strings, and for identifying the most suitable search engines. The search string below has been used in each search, after adaptation to each search engine.

[title, abstract] (((“eco-anxiety” OR “eco anxiety” OR (“eco*” W0 (“trauma” OR “stress” OR “anxiety” OR “grief” OR “guilt” OR “coping” OR “depression” OR “anger” OR “distress“)) OR (“climate” W0 (“worry” OR “anxiety” OR “stress” OR “concern” OR “uncertainty” OR “distress“)) OR (“climate change” W0 (“worry” OR “anxiety” OR “stress” OR “concern” OR “uncertainty” OR “distress“)) OR (“environmental” W0 (“worry” OR “anxiety” OR “stress” OR “concern” OR “distress“)) OR (“global warming” W0 (“worry” OR “anxiety” OR “stress” OR “concern” OR “uncertainty“)) OR “solastalgia“)) AND (“test*” OR “scale” OR “measure*” OR “tool” OR “psychometric” OR “questionnaire” OR “assess*” OR “inventory” OR “index“).

### Selection Criteria

Original articles describing the psychometric properties and/or validity of self-report measures designed to assess eco-anxiety and other climate change-related emotions in the general population were considered for inclusion. Studies were not restricted in terms of country, age, race, gender, or publication language. The following exclusion criteria were used: (1) papers reporting tools designed to assess institutionalized, inpatient, or psychopathological samples; (2) articles focused on behavioural or cognitive reactions to the climate crisis rather than emotions; (3) abstract-only papers, under review papers, preprints, conference materials, editorials, author responses, theses, or books; (4) articles reporting tools for qualitative data collection; and (5) reviews, meta-analyses, or commentaries.

### Review Process

The searches were conducted on the 6^th^ of September 2023 in each search engine separately: PubMed, PsycINFO, and Web of Science. Compared to the initial protocol, the Scopus search engine was replaced with Web of Science because in Scopus the search string as it was devised was yielding to an unexpected high volume of items.

The full list of articles retrieved from the three search engines was saved in Rayyan,^33^ eliminating duplicates. All titles and abstracts were reviewed by one researcher (FKA), irrelevant items were discarded. Full text for the potentially eligible items was downloaded, and inclusion and exclusion criteria applied blindly by two reviewers (FKA and MJC). No discrepancy between reviewers was reported. Articles reporting behavioural and cognitive reaction to the climate crisis^34^ rather than measuring emotions, articles reporting instruments to measure a more generic worry not only towards climate change but also various risks that might damage nature, such as potential nuclear accidents,^35^ and articles reporting instruments targeting environmental concern only (cognitive reaction) were excluded. The emotional component of environmental concern was conversely included when assessed together with other emotions. Finally, studies including only one question to assess to what extent people were worried, anxious, depressed, or angry about global warming or climate change^11,36^ without following any scale development procedure, were excluded. Subsequently, the reference lists of the included articles were searched to identify further studies. Numbers of included and excluded articles were recorded at each stage using the PRISMA diagram.^32^

For each included article describing a new psychometric instrument information was extracted on an excel spreadsheet: definition of the eco-emotion used, country and language in which the scale was developed, title of subscales (if any), example items for (sub)scales, and emotion terms in the scale and their frequency of use. For the articles reporting validation of existing instruments, the following data were extracted: country and language in which the study was conducted, number of items, factorial structure of the scales, types and results of factor analyses, features of target population, age and gender of the samples, internal consistency values of (sub)scales, sample size, gender differences, variables used to test the validity of the scales, and results of analyses to test validity. Data extraction was carried out by one author (FKA) and the results were checked by another (MJC), independently. One author (NH) checked the reference lists of the existing articles and extracted and listed the emotions included in the scales.

To rigorously appraise the study designs and risk of bias of the included articles, the COSMIN Study Design^37^ and COSMIN Risk of Bias (RoB)^31^ checklists were employed. Studies were appraised against the criteria in the checklists, adopting the following scoring rubric: ‘very good’ (3), ‘adequate’ (2), ‘doubtful’ (1), ‘inadequate’ (0), and ‘not applicable’ (0). For the Patient-Reported Outcome Measures (PROM) development section, a distinction was made for adaptation studies which needed to satisfy general design prerequisites only, and development studies evaluated against the entire suit of criteria. Scores were allocated according to the study’s adherence to the COSMIN criteria and then summed across each category, leading to a composite score for each domain. For the Risk of Bias assessment, the same score delineated whether the study bore a high, moderate, or low risk of bias, according to the performance across the checklist domains. In setting the scores from each COSMIN domain, we established threshold values to ascertain the risk of bias and the design quality for each study. A high risk of bias was attributed to studies with low scores in more than two COSMIN domains; low scores in two domains led to a moderate risk, while a low risk was attributed to those with low scores in a single domain. In terms of design quality, studies without any low scores were judged to have good quality; low scores in one or two domains led to adequate quality, and those with three or more low scores were labelled doubtful.

## Results

Of the 55 instrument evaluated, 17 met the inclusion/exclusion criteria. A further 5 studies were identified through citation searching leading to a total of 22 papers included in this systematic review (Figure 1). Ten studies described the development of psychometric scales measuring various eco-emotions; the remaining twelve studies were validation of some of these scales in different samples.

**Figure 1.**
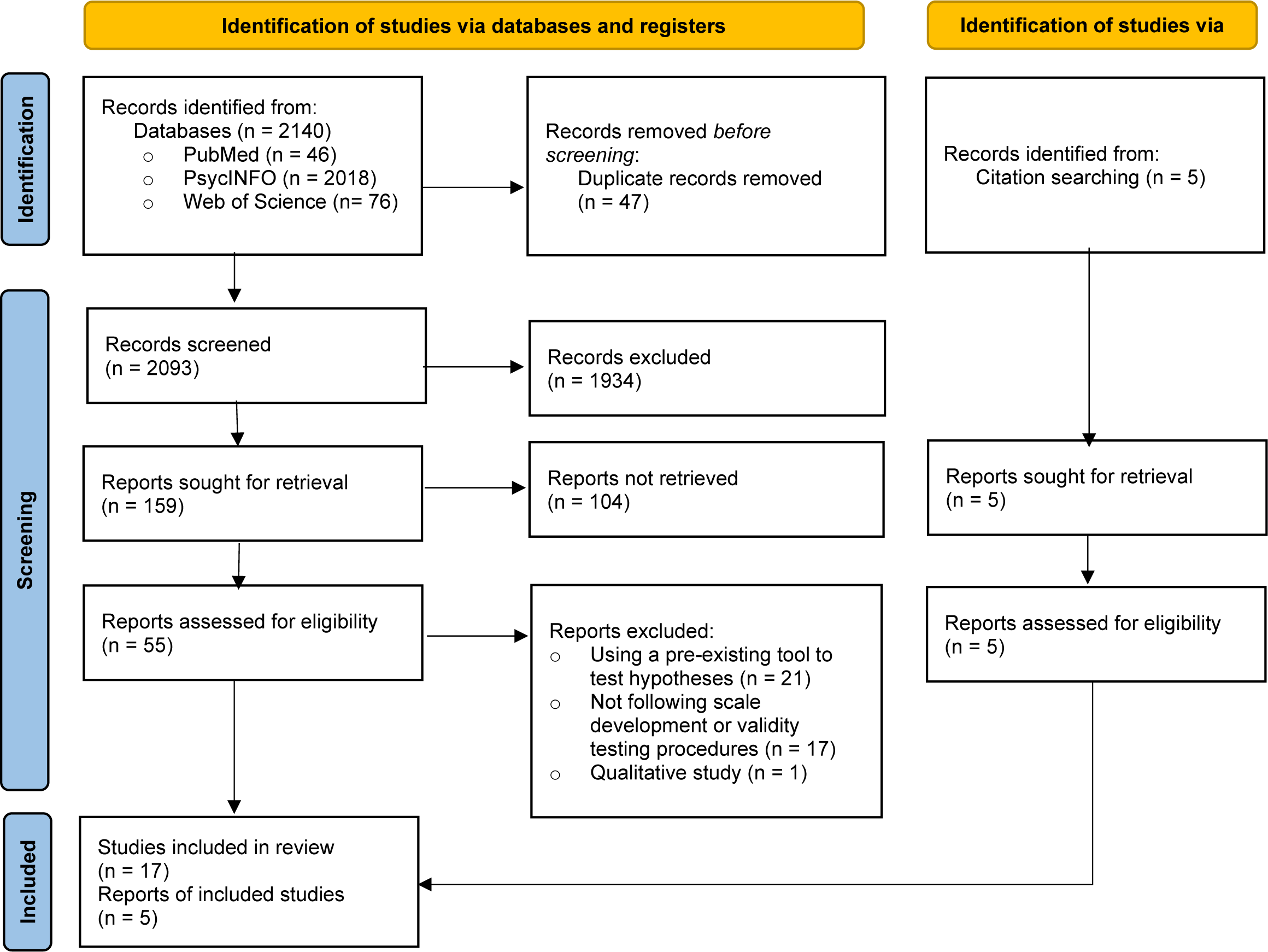
A flow chart of systematic review based on PRISMA (Page et al., 2020)

### General characteristics

The general characteristics of the instruments, including the emotions assessed, and their definitions are reported in Table 1. One instrument measured environmental anxiety,^38^ two eco-anxiety,^13,17^ one climate change anxiety.^15^ The other emotions studied included climate change worry,^39^ climate change distress,^40^ solastalgia,^41^ solastalgia as a subset of environmental distress,^16^ eco-guilt,^13^ and eco-grief.^13^

**Table 1.** General Characteristics of the Scales.

| Name of the Scale<br>Source | Name of the<br>Concept | Definition of the Concept | Country /<br>Language | Subscale(s) | Example item(s) | Emotions<br>(frequencies) |
| --- | --- | --- | --- | --- | --- | --- |
| **Environmental**<br>**Worry Scale**<br>**Bowler &**<br>**Schwarzer, 1991** | Environmental<br>anxiety | "An emotion towards industrial or<br>environmental risks" (p. 168) | USA /<br>English | – | "I get easily upset when thinking<br>about poisons in my<br>environment" | Worry (3)<br>Fear (2)<br>Upset (1) |
| **Environmental**<br>**Distress Scale**<br>**Higginbotham et**<br>**al., 2006** | Solastalgia | "The sense of distress people<br>experience when valued environments<br>are negatively transformed" (p. 245) | Australia /<br>English | Solastalgia*,<br>Frequency,<br>Observation,<br>Threat, Impact,<br>Actions | "I miss having the sense of peace<br>and quiet I once enjoyed in this<br>place" | Sadness (2)<br>Worry (1)<br>Shame (1)<br>Upset (1) |
| **Climate Change**<br>**Distress Scale**<br>**Searle & Gow,**<br>**2010** | Climate change<br>distress | "Intense worry that leads to distress<br>and/or interferes with daily living" (p.<br>362) | Australia /<br>English | 1-Climate change<br>anxiety<br>2-Climate change<br>hopelessness | "Thinking about climate change<br>now makes me feel ..." rated for<br>each emotion. | Concern (1)<br>Tension (1)<br>Worry (1)<br>Anxiety (1)<br>Depression (1)<br>Hopelessness (1)<br>Powerlessness (1)<br>Sadness (1)<br>Helplessness (1)<br>Stress (1)<br>Anger (1)<br>Fear (1) |
| **Climate Change**<br>**Anxiety Scale**<br>**(CCAS)**<br>**Clayton &**<br>**Karazsia, 2020** | Climate change<br>anxiety | "Clinically significant response toward<br>climate change" (p. 9) | USA /<br>English | 1-Cognitive-emotional<br>impairment,<br>2-Behavioural<br>engagement, 3-Experience,<br>4-Functional<br>impairment | S1: "Thinking about climate<br>change makes it difficult for me<br>to concentrate" S2: "I feel guilty<br>if I waste energy"<br>S3: "I know someone who has<br>been directly affected by climate<br>change"<br>S4: "My concerns about climate<br>change make it hard for me to<br>have fun with my family or<br>friends" | Concern (4)<br>Guilt (1)** |
| **Hogg Eco-Anxiety Scale (HEAS)**<br>**Hogg, et al., 2021** | Eco-anxiety | "Experiences of anxiety relating to environmental crises. ...It encompasses "climate change anxiety" ... as well as anxiety about a multiplicity of environmental calamities, which may or may not be directly caused by climate change." (p. 1) | New Zealand / English | 1-Affective symptoms,<br>2-Rumination, 3-Behavioural symptoms,<br>4-Personal impact anxiety | S1 "Feeling nervous, anxious or on edge" S2: "Unable to stop thinking about past events related to climate change"<br>S3: "Difficulty working and/or studying"<br>S4: "Feeling anxious about the impact of your personal behaviours on the earth" | Anxiety (4)<br>Worry (2)<br>Fear (1)<br>Nervousness (1) |
| **Climate Change Worry Scale**<br>**Stewart, 2021** | Climate change worry | "Verbal-linguistic thoughts about the changes that may occur in the climate system and the possible effects of these changes" (p. 1) | USA / English | – | "I worry about climate change more than other people." | Worry (9) |
| **Eco-Anxiety Questionnaire**<br>**Agoston et al., 2022** | Eco-anxiety | "A chronic fear or non-specific worry of environmental doom" (p. 2) | Hungary / Hungarian | 1-Habitual ecological worry,<br>2-Negative consequences of anxiety | S1: "I am worried about the increasing number of natural disasters caused by climate change"<br>S2: "I am so anxious about climate change that it affects my performance at school/work" | Anxiety (2)<br>Worry (3)<br>Fear (3)<br>Anger (2)<br>Uneasiness (1)<br>Upset(2) |
| **Eco-Guilt Questionnaire**<br>**Agoston et al., 2022** | Eco-guilt | "A specific form of guilt that people experience when they feel they are not meeting personal or societal environmental standards, or when they contemplate polluting activities" (p. 3) | Hungary / Hungarian | – | "The more I know about the human causes of climate change, the more things I feel guilty about" | Guilt (4)<br>Anger (1)<br>Uneasiness (1) |
| **Ecological Grief Questionnaire**<br>**Agoston et al., 2022** | Eco-grief | "Grief experienced in relation to anticipated or experienced ecological loss due to acute or chronic environmental change, including loss of species, ecosystems, or beloved landscapes" (p. 2) | Hungary / Hungarian | – | "It is frightening that climate change is causing the destruction of natural areas at such a dramatic rate that they will never be the same again." | Sadness (1)<br>Fear (1) |
| **Scale of Solastalgia** | Solastalgia | "The distress caused by a change in an appreciated place and its cumulative impact on the mental health of those | Chile / not reported | 1-Solace,<br>2-Algia | S1: "It makes me sad to think that one day I will be forced to | Sadness (3)<br>Distress (1)<br>Anxiety (1) |
| **Caceres et al.,**<br>**2022** |  | who live in that specific location" (p. 1) |  |  | leave this place as a result of climate change."<br>S2: "Lately, I feel anxious." | Worry (1)<br>Fear (1)<br>Tension (1)<br>Nostalgia (1) |

*Note.* The scales were sorted based on the publication dates of the studies. The last column indicates the emotions or affects mentioned in each scale and the number of items mentioning these emotions or affects.
\* The solastalgia subscale of the Environmental Distress Scale was taken into account in this review based on the scope of the current review.
\*\* The item including the feeling of "guilt" was not included in subsequent validation studies because it belongs to the behavioural engagement subscale which was not involved in 13-item version of the CCAS.

### Definitions and frequency of emotions

Among the studies measuring anxiety, the only study^38^ published before the APA’s report^6^ defined the concept as a sentiment regarding potential hazards in industry or the environment. In the remaining studies, one paper only^13^ conformed to the APA’s definition for eco-anxiety, while the other two instruments used different definitions^15,17^ (Table 1).

Considering that some emotions are tightly linked one to another (e.g. worry and anxiety), we mapped the constellation of emotions assessed by each individual tool, regardless of the overarching aim of the tool itself. For example, the Eco-Anxiety Questionnaire^13^ aimed to assess eco-anxiety, however a wider number of tightly related negative emotions were also assessed, i.e. worry, fear, and upset (Table 1). The most recurrent emotion was worry which was used twenty times, followed by fear (nine times), anxiety (eight times), sadness (seven times), concern (five times), anger, and upset (four times), uneasiness and tension (twice), and nervousness, stress, distress, shame, depression, hopelessness, powerlessness, helplessness, and nostalgia used only once. Instead of developing situation-specific statements, Searle and Gow^40^ designed the Climate Change Distress scale (CCDS) to ask participants to rate twelve emotions from a list; thus this scale presented the greatest variety of emotions. However, the reason why all emotions were gathered under the umbrella term of “distress” was not explained (Table 1).

Since the 13-item version of CCAS^15^ did not contain behavioural engagement and experience of climate change subscales, one item containing the feeling of “guilt” which belongs to the behavioural engagement subscale, was not tested in the subsequent validation studies of 13-item CCAS. Only the items containing “concern” were employed in the validation studies of CCAS.^42–49^ One scale only included worry as the single type of emotion, used nine times.^39^ All but one scale^40^ consists of statements aimed at measuring emotions, thoughts and/or behaviours regarding climate crisis or environmental changes due to climate crises.

### Language and setting

Six of the published instruments were developed in English^15–17,38–40^ while three of them (in the same article) were developed in Hungarian.^13^ One article did not specify the language of the instrument but it is likely to be Spanish considering that it was tested in Chile.^41^

The instruments were tested in different countries: three in the United States,^15,38,39^ two in Australia,^16,40^ three in Hungary,^13^ one in New Zealand,^17^ and one in Chile.^41^ Ágoston et al.^13^ administered the eco-anxiety, eco-guilt, and eco-grief scales to the same participants in Hungary.

Two of the original scales^17,39^ employed university students for their development. Three studies collected data from people living in areas exposed to toxicity^38^, environmental disturbances (i.e., open-cut mining area^41^), or natural disasters.^16^ Two of them compared these data with those collected from people who live in areas not directly exposed to environmental deterioration,^38,41^ indicating the discriminant validity of the tools.

### Psychometric characteristics and validation data

Psychometric characteristics of the ten included instruments and of all the 22 validation studies identified from the search were summarized Table 2. Here, of the ten instruments included in Table 1, only two (i.e. CCAS^15^ and Hogg Eco-Anxiety Scale (HEAS)^17^) were validated on external populations (a total of nine studies reported the validation of the CCAS, two of the HEAS, and one compared them both). Factor structures, and validity results of all scales are presented in Supplementary Material 1.

**Table 2.** Findings from the studies testing the validity of the scales.

| Name of the scale | Source | Country / Language | Number of items | Number and names of factors (number of items per factor) / analysis type / sample size | Target population Age and Gender | (Sub)scale Reliability (Cronbach's $\alpha$) | Gender (Female/Male) Differences and analysis type | Variables used to gather validity evidence on test relationships |
| --- | --- | --- | --- | --- | --- | --- | --- | --- |
| Environmental Worry Scale (EWS) (Scale development) | Bowler & Schwarzer (1991) | USA / English | 17 and 8 (for short version) | one-factor (environmental worry)<br>EFA or CFA not conducted<br>n = 547 (pretested in a sample of 250 undergrads) | General population in USA (Those exposed to and those distant from the toxic areas)<br>no info about age or gender | 0.77 for 8-item | No difference correlation | Tension, depression, anger, vigor, fatigue, confusion, anxiety, exposure |
| Environmental Distress Scale (Solastalgia subscale) * (Scale development) | Higginbotham et al. (2007) | Australia / English | 9 | one-factor for solastalgia (9)<br>EFA using PCA one-factor solution for solastalgia subscale<br>n = 203 | general population in Australia (Those exposed to environmental disturbances ($M_{age} = 48$, 48% female) and not exposed to environmental disturbances ($M_{age} = 54$, 46% female)) | 0.93 | No difference t-test | Comparison of non-exposed vs. exposed groups |
| Climate Change Distress Scale (Scale development) | Searle & Gow (2010) | Australia / English | 12 | two-factor (climate change anxiety (9), climate change hopelessness (3)) | general population in Australia<br>no info. about age or gender | 0.92 for climate change anxiety, 0.82 for climate change hopelessness | F > M in climate change anxiety<br>t-test | Environmental beliefs, future anxiety, intolerance of uncertainty, religiosity |
|  |  |  |  | EFA using PCA with oblique rotation<br>n = 275 |  |  |  |  |
| Climate Change Anxiety Scale (CCAS) (Scale development) | Clayton & Karazsia (2020) | USA / English | 22 | four-factor (cognitive-emotional impairment (8), behavioural engagement (6), experience (3), functional impairment (5))<br>EFA using PAF with oblimin rotation suggesting four factor solution<br>n=197<br>CFA<br>n=199 | general population in USA<br>50% were between age 25 and 34, range: 18-84<br>%48 female | 0.97 for cognitive-emotional impairment, 0.79 for behavioural engagement, 0.86 for experience, 0.94 for functional impairment | F > M in behavioural engagement t-test | Environmental identity, negative emotionality, depression/anxiety |
| CCAS (Scale validation) | Innocenti et al. (2021) | Italy / Italian | 13 | two-factor used (cognitive-emotional impairment (8), functional impairment (5))<br>EFA recommended one-factor<br>n = 150<br>CFA to test two-factor model | Italian adults<br>M<sub>age</sub> = 32.84 (SD = 11.72), range: 19-76<br>67% female | 0.78 for cognitive-emotional impairment, 0.72 for functional impairment | Not reported | General anxiety disorder, distress, depression, anxiety, new social paradigm, self-efficacy, dominant paradigm, pro-environmental behaviours |
| CCAS<br>(Scale validation) | Wullenkord et al. (2021) | Germany / German | 13 | one-factor (climate change anxiety)<br>CFA<br>n = 1011 | general population in German<br>M<sub>age</sub> = 43.91 (SD = 13.97), range: 18-69<br>51% female | 0.89 | F > M in overall climate change anxiety t-test | Pro-environmental intentions, avoidance, denial of personal outcome severity of climate change, human dominance over nature, general anxiety and depression, competence frustration right-wing political orientation, denial of guilt |
| CCAS<br>(Scale validation) | Cruz & High (2022) | USA / English | 13 (does not fit) (11-item restructured version best fit) | one-factor (climate anxiety)<br>CFA<br>n = 513 | general population in USA<br>M<sub>age</sub> = 52.20 (SD = 18.54)<br>%61 female | NR | Not reported | Depression, trait anxiety, state anxiety |
| CCAS<br>(Scale validation) | Larionow (2022) | Poland / Polish | 13 | three-factor (functional impairment (5), intrusive symptoms (4), reflections on climate anxiety (4))<br>EFA with oblimin rotation with 3-factor solution<br>CFA<br>n = 603 | Polish adults<br>M<sub>age</sub> = 25.32 (SD = 9.59), range: 18-70<br>57% female | 0.89 for functional impairment, 0.83 for intrusive symptoms, 0.77 for reflections on climate anxiety | F > M in cognitive impairment and functional impairment Mann-Whitney U-test | Experience of climate change, behavioural engagement, environmental identity, biospheric concerns, altruistic concerns, egoistic concerns, climate change denial, anxiety symptoms, depressive symptoms, anxiety-depressive symptoms, sense of safety, self-blame, acceptance, rumination, positive refocusing, refocus on planning, positive reappraisal, putting into perspective, catastrophizing, blaming others |
| CCAS (Scale validation) | Mouguiama-Daouda et al. (2022) | France / French | 13 | two-factor (cognitive-emotional impairment (8), functional impairment (5))<br>CFA<br>n = 305, n = 905 | general population in France<br>M<sub>age</sub> = 30.80 (SD = 11.32)<br>72% female | 0.84 for cognitive-emotional impairment, 0.82 for functional impairment | Not reported | Depression, general anxiety disorder, environmental identity |
| CCAS (Scale validation) | Simon et al. (2022) | Philippines / English | 13 | two-factor (cognitive-emotional impairment (8), functional impairment(5))<br>CFA<br>n = 452 | Filipino adolescents (undergraduates)<br>M<sub>age</sub> = 19.18 (SD = .99)<br>no info. for gender | 0.90 for cognitive-emotional impairment, 0.85 for functional impairment | NR | Validity test based on computations of composite reliability (CR), average variance extracted (AVE), and maximum shared variance (MSV) |
| CCAS (Scale validation) | Jang et al. (2023) | Korea / Korean | 13 | two-factor (cognitive-emotional impairment (8), functional impairment (5))<br>EFA using PCA with varimax orthogonal rotation<br>n = randomly selected 350 out of 459 | Korean adults<br>M<sub>age</sub> = 44.18 (SD = 13.54), range: 19-65<br>51% female | 0.85 for cognitive-emotional impairment, 0.89 for functional impairment | NR | Validity test based on computations of composite reliability (CR) and average variance extracted (AVE) |
|  |  |  |  | CFA<br>n = 459 |  |  |  |  |
| CCAS<br>(Scale<br>validation) | Tam et al.<br>(2023) | China, India,<br>Japan, and<br>USA / English,<br>Chinese,<br>Japanese | 13 | two-factor<br>(cognitive-emotional<br>impairment (8),<br>functional<br>impairment (5))<br>CFA two-factor<br>N = 4000 (1000<br>from each<br>country) | general population<br>(age over 18)<br>percentages<br>reported for age<br>ranges<br>49% female in<br>China and India,<br>52% in Japan and in<br>USA | 0.93, 0.88,<br>0.93, 0.95 for<br>cognitive-emotional<br>impairment,<br>0.91, 0.87,<br>0.93, 0.94 for<br>functional<br>impairment by<br>country ranking | M > F in<br>cognitive<br>impairment<br>and functional<br>impairment in<br>China and USA<br>correlation | Climate change belief<br>in happening, climate<br>change belief in<br>scientific consensus,<br>worry, perceived harm<br>to self, perceived harm<br>to country, climate<br>action, resource<br>conservation,<br>sustainable diet,<br>climate activism,<br>support for climate<br>policy |
| CCAS<br>(Scale<br>validation) | Wu et al.<br>(2023) | Canada /<br>English | 4 | one-factor<br>(climate anxiety)<br>EFA<br>n = 1144<br>CFA<br>n = 1162 | Canadian<br>adolescents<br>M<sub>age</sub> = 16.3 (SD =<br>0.5), range: 15-18<br>45% female | 0.95 | NR | general anxiety,<br>depression, climate<br>concern, positive<br>mental health, life<br>satisfaction, self-<br>concept |
| Hogg Eco-Anxiety Scale<br>(HEAS)<br>(Scale<br>development) | Hogg et al.<br>(2021) | New Zealand<br>/ English | 13 | four-factor<br>(affective<br>symptoms (4),<br>rumination (3),<br>behavioural<br>symptoms (3),<br>personal impact<br>anxiety (3))<br>EFA using PCA<br>with oblimin<br>rotation<br>n = 343 | undergraduate<br>students<br>sample one: M<sub>age</sub> =<br>19.90 (SD = 3.59);<br>79.7% female<br>sample two: M<sub>age</sub> =<br>19.42 (SD = 2.87);<br>69% female | 0.92 for<br>affective<br>symptoms, 0.90<br>for rumination,<br>0.86 for<br>behavioural<br>symptoms, 0.88<br>for personal<br>impact anxiety | NR | Stress, anxiety,<br>depression, emotional<br>reactivity, credibility of<br>science, climate change<br>belief |
|  |  |  |  | CFA<br>n = 342 |  |  |  |  |
| HEAS<br>(Scale<br>validation) | Uzun et al.,<br>2022 | Türkiye/Turkish | 13 | four-factor<br>(affective<br>symptoms (4),<br>rumination (3),<br>behavioural<br>symptoms (3),<br>personal impact<br>anxiety (3))<br>EFA by using PCA<br>n = 698<br>CFA | general population<br>(over age 18)<br>M<sub>age</sub> = 23.07 (SD =<br>6.01)<br>72% female | 0.83 for<br>affective<br>symptoms, 0.84<br>for rumination,<br>0.86 for<br>behavioural<br>symptoms, 0.84<br>for personal<br>impact anxiety | NR | Validity test based on<br>computations of<br>composite reliability<br>(CR) and average<br>variance extracted<br>(AVE) |
| HEAS<br>(Scale<br>validation) | Pavani et al.,<br>2023 | France /<br>French | 13 | one-factor<br>(eco-anxiety)<br>EFA or CFA not<br>conducted<br>n = 200 | general population<br>(over age 18)<br>M<sub>age</sub> = 32.42 (SD =<br>14.75)<br>71% female | 0.94 | NR | pro-environmental<br>behaviours |
| CCAS and<br>HEAS<br>(Scale<br>validation) | Hogg et al.,<br>2023 | Australia /<br>English | 22 for<br>CCAS &<br>13 for<br>HEAS | four-factor<br>(affective<br>symptoms (4),<br>rumination (3),<br>behavioural<br>symptoms (3),<br>personal impact<br>anxiety (3)) for<br>HEAS and two-factor (cognitive-emotional<br>impairment (8),<br>functional<br>impairment (5)) | general population<br>in Australia<br>M<sub>age</sub> = 39.49 (SD =<br>16.46), range: 18-86<br>63% female | 0.88 for<br>affective<br>symptoms, 0.86<br>for rumination,<br>0.78 for<br>behavioural<br>symptoms, 0.86<br>for personal<br>impact anxiety<br>(HEAS); 0.85 for<br>cognitive-emotional<br>impairment,<br>0.83 for<br>functional | F > M in<br>affective<br>symptoms and<br>personal<br>impact anxiety<br>t-test | risk perception, direct<br>experience,<br>information seeking,<br>information avoidance |
|  |  |  |  | for CCAS with 13-item Multigroup CFA n = 530 |  | impairment (CCAS) |  |  |
| Eco-Anxiety Questionnaire (Scale development) | Agoston et al., 2022 | Hungary / Hungarian | 22 | two-factor (habitual ecological worry (13), negative consequences of anxiety (9))<br>EFA with WLSMV estimation and geomin rotation n = 1152 out of 4608<br>CFA with WLSMV estimation n = 1152 out of 4608 | adults<br>M<sub>age</sub> = 43.3 (SD = 13.2)<br>41% female | 0.91 for habitual ecological worry, 0.86 for negative consequences of anxiety | F > M in habitual ecological worry and negative consequences of anxiety t-test | Pro-environmental behaviours |
| Eco-Guilt Questionnaire (Scale development) | Agoston et al., 2022 | Hungary / Hungarian | 11 | one-factor (eco-guilt)<br>EFA with WLSMV estimation and geomin rotation n = 1152 out of 4608<br>CFA with WLSMV estimation n = 1152 out of 4608 | adults<br>M<sub>age</sub> = 43.3 (SD = 13.2)<br>41% female | 0.89 | F > M in eco-guilt t-test | Pro-environmental behaviours |
| Ecological Grief Questionnaire (Scale development) | Agoston et al., 2022 | Hungary / Hungarian | 6 | one-factor (ecological grief)<br>EFA with WLSMV estimation and geomin rotation<br>n = 1152 out of 4608<br>CFA with WLSMV estimation<br>n = 1152 out of 4608 | adults<br>M<sub>age</sub> = 43.3 (SD = 13.2)<br>41% female | 0.77 | F > M in ecological grief t-test | Pro-environmental behaviours |
| Climate Change Worry Scale (Scale development) | Stewart, 2021 | USA / English | 10 | one-factor (climate change worry)<br>EFA<br>n = 600<br>CFA | university students<br>study 1: M<sub>age</sub> = 22.3 (SD = 5.9), range: 18-51, %50 female;<br>study 2: M<sub>age</sub> = 20.9 (SD = 1.06), 83% female; study 3: M = 20.8 (SD = 1.9), range: 18-37<br>85% female | 0.95 | No difference invariance analysis | Political orientation, fear of weather, storm fear, stress, anxiety, depression, worry |
| Scale of Solastalgia (Scale development) | Caceres et al., 2022 | Chile / no info. | 10 | two-factor (solace (7), algia (3))<br>EFA by using PFA with oblimin rotation<br>n = 223 | general population living in areas exposed to forest fires and droughts (over age 18)<br>no info. about age<br>58% female | 0.87 for solace, 0.90 for algia | NR | Post-traumatic stress disorder |

Note. NR: Not reported, PCA: Principal Component Analysis, PFA: Principal Factor Analysis.
\* The solastalgia subscale of the Environmental Distress Scale was involved in this review based on the scope of the current review

### Dimensionality

Six instruments were multidimensional;^13,15–17,40,41^ four unidimensional.^13,38,39^ The Environmental Distress Scale^16^ originally consists of six subscales. Only the subscale assessing solastalgia was included in this review as the other five subscales included the frequency of environmental threats in the area people live in and other contextual characteristics and have not met the inclusion criteria of this review.

Different versions of the same multidimensional scale were published: in the original paper describing the CCAS,^15^ Clayton and Karazsia presented a 13-item scale claiming that the climate anxiety response was composed of two factors: cognitive-emotional impairment and functional impairment. However, the authors did not report model fit statistics for this two-factor scale; the model fit for a four-factor model (22 items including also ‘pro-environmental behaviour’ and ‘experience of climate change’) was instead reported. Moreover, the psychometric performance as a one-factor model was also not reported, making it unclear if the scale, as one-dimension, can be used to measure climate anxiety.

### Behavioural, cognitive and physical components

As a result of the different conceptualization of eco-emotions addressed in the studies, the comprehensiveness and dimensions of the measurement tools also changed. While all the instruments kept the focus of their assessment on the climate-related emotion, some of them added the assessment of behavioural or cognitive or physical aspects related to the underlying emotion. All but one scale^40^ included at least one item indicating behavioural (e.g., sleeping poorly because of thinking about climate change^15^), cognitive (e.g., being unable to stop thinking about past events related to climate change^17^), or physical reactions (e.g., experiencing unusual tension in muscles^13^) toward climate crises. Some instruments also included items representing the experience about climate change (e.g., life threatening water scarcity^41^), or observation of the environmental changes (e.g., change of the wildlife around the person in a disturbing way^13^) (Table 1).

### Validation in additional populations

The most analysed scale in terms of psychometric properties was the CCAS,^15^ however the findings differed in terms of target sample, dimensionality of the scale, and the number of items used. The validity of CCAS was assessed among both adolescents^47,50^ and adults.^15,44–46,48,49,51,52^ The validation studies of CCAS used either the 22-item or 13-item versions of the scale (full description available in the Supplementary Material 1). The validation of HEAS with 13-item was tested in four countries: New Zealand,^17^ Türkiye,^53^ France,^54^ and Australia.^51^ Four-factor structure was supported by three out of four studies; however, a one-factor solution showed better fit in another study.^54^ One study employed both CCAS and HEAS to statistically compare their psychometric properties in the same sample.^51^ The subscales of the two instruments were found to be highly and positively correlated with each other (correlation values ranged from 0.58 to 0.69).

All but one^42^ studies calculated Cronbach’s alpha as a measure of scale reliability. The internal consistency of the subscales ranged from 0.72 to 0.97 (Table 2). Three of the studies reported test-retest reliability.^16,17,39^ A preliminary study (pretest of the scale with another sample or thematic analysis of interviews) was conducted by four studies before developing^13,16,38^ or adapting a scale.^54^

### Gender differences

Ten studies did not report whether they tested for possible gender differences. Three of the studies found no gender difference in terms of the construct they tested.^16,38,39^ Among the participants filling out the CCDS, women were found to have higher levels of climate change anxiety and climate change hopelessness than men.^40^ A gender difference was not found in the CCAS^15^ in terms of cognitive-emotional impairment or functional impairment, but for behavioural engagement women reported higher levels than men. In Wullenkord,^49^ compared to men, women reported higher levels of climate change anxiety (overall score); while Larionow^45^ found that women had higher cognitive impairment and functional impairment in CCAS. Testing the validation of CCAS in four countries, Tam^48^ revealed that men in China and the US had higher levels of cognitive impairment and functional impairment, as compared to women. Hogg^17^ did not find a gender difference in CCAS, but using HEAS found that women experienced higher levels of affective symptoms and personal impact of anxiety. Ágoston^13^ stated that women experienced greater eco-guilt, eco-grief, and habitual ecological worry and negative consequences of anxiety indicating higher levels of eco-anxiety.

### COSMIN Evaluation

One of the scale development studies scored “good”^17^ and one had “doubtful”^38^ quality ratings in terms of study design. One of the validation studies of CCAS scored “good”,^52^ one of the validation studies of HEAS “doubtful” quality.^53^ The remaining studies developing or validating a scale had “adequate” quality. In addition, all but one studies developing an instrument were rated to have moderate level of risk of bias. Bowler and Schwarzer’s study^38^ had high risk of bias. Four validation studies of CCAS^45,45–47,49^ had moderate risk of bias. One validation study of HEAS had moderate^54^ and one had high risk of bias.^53^ The risk of bias of other studies was rated as low. The tables of assessment results were presented as a Supplementary Material 2.

## Discussion

This is the first systematic review mapping the psychometric characteristics of instruments assessing eco-emotions and their validation in different populations. A total of ten instruments were reviewed covering a wide range of negative emotions including anxiety, distress, worry, guilt, grief, and solastalgia. Two of these instruments have been also validated in other populations.

A preliminary study to define the range of eco-emotions to be potentially included to provide content validity evidence and test-retest reliability of the measures were missing in most of the included scales, despite both components have been emphasized to be relevant prerequisites for scale development.^30,55–57^ According to the assessments of study design and risk of bias of each study, most studies seem in need of improvement and better evidence in terms of methodological robustness, and to be at moderate risk of bias.

This review provides a map of which emotions were included in the scales and their frequency of use. Although the importance of addressing the positive emotions in this context has been emphasized,^13,23^ no positive emotions were included in any of the scales. ‘Worry’ - not ‘anxiety’ - was the most recurrent emotion; these show common features but differ in terms of the realistic concern and duration of symptoms. Worry is based on concrete situations and usually temporary, but when it persists in a person’s life and is accompanied by increased autonomic arousal, it can be related to other negative emotional and cognitive experiences, such as generalized anxiety and rumination.^25,58^ Although worry and anxiety have conceptual differences, respondents may struggle to accurately identify the psychological reactions they have to various potential impacts of climate change.^59^ While the use of ‘anxiety’ instead of ‘worry’ and vice versa may seem like a deliberate choice, there is no explicit mention of the concrete difference between them or that they are interchangeable structures. Thus, to what extent these emotions may overlap or vary in relation to climate change still needs to be clarified. This is partly due to the lack of consistent definitions of climate-related emotions in scale development studies.

The studies included in this review use various definitions of emotions, not always referring to the APA definition of eco-anxiety.^6^ This is probably because this definition is nonetheless vague and not specific enough to allow a strict adherence to the underlying concept. Moreover, there is also a lack of consistency of assessment of the dimensions of each emotion as seen in the predominance of affective symptoms^40^ but also cognitive, physiological, and behavioural indicators of eco-anxiety.^17^ Exploring these different components of emotions would increase the depth of the assessment, although this needs to be counterbalanced with the length of the instrument. Therefore, it is difficult to assess whether all these studies examine the same concept and measure the same structure.

Researchers involved in the development of scales to measure eco-emotions emphasize the necessity of generating a clear definition of the concept^15^ but none of them has provided a new and concrete definition according to the scale they have developed and the data they have collected. In addition, there is little clarity also on the definition of the ecological component of eco-emotions. Currently there is no consensus in the literature as to whether it is necessary to address only concern about human-induced damages to nature or to measure concern for all changes in the environment.^55^ As a consequence, some authors^13^ refer uniquely to climate change-related events, while others^38^ also include man-made disasters that might have environmental consequences (i.e. nuclear power accidents). The reviewed instruments can be regarded as preparatory work to develop a theoretical framework to assess the mental health consequences of the climate crisis, at population level. This must include not only clear definitions of the emotions involved (including positive emotions), and a characterisation of their dimensions (cognitive, behavioural, physical, etc.), but also a solid reference to the context that elicits them. In this respect we advocate for using a Planetary Health approach by considering the impacts of human disruption on Earth’s natural system on human health and all life on Earth.^60^

Cultural and geographical variations remain a withstanding challenge in this field. Not only the effects of climate change are geographically unequally distributed and responsible for unjust disruption of human and ecosystem health, but the intimate relationship between humans and the surrounding natural environment also changes across cultures^1^. For example, farmers living in regions experiencing severe droughts due to climate change face challenges like water scarcity and crop failures, placing a disproportionate burden on their livelihoods. At the same time, in low-lying regions where increased rainfall and rising sea levels are contributing to more frequent and severe floods, communities try to implement strategies such as rainwater harvesting systems and improved drainage infrastructure to manage excess water during heavy rainfall. The cross-cultural invariance of CCAS, the cross-cultural validity of an instrument assessing eco-emotions from wider perspective in this context becomes extremely relevant.^48^

In this review, six of the scales were originally developed in English and validated in Western, educated, industrialized, rich, and democratic (WEIRD) populations, despite these representing only 12% of the world population.^61^ The validation of four out of six measures was not tested in another country or language. However, it is emphasized in the current literature that presumptions derived from Western contexts may not consistently function in the same way when applied to non-Western environments.^62^ This calls for the need for cultural adaptation of given instruments. Nonetheless, the cultural adaptation of existing instruments is also a function of the definition of the environmental/climate crisis in relation to eco-emotions as some consequences of climate change might have a very different relevance to geographically distant populations. For example, temperature warming might be perceived as pleasant in some Nordic area,^63^ or the emotion eco-guilt may be more relevant in more urban, industrialized (emission outputting) cultural contexts.

Not only were the emotions studied usually vaguely defined, but their own mention derived by a top-down approach by identifying the key components of the specific phenomenon of interest based on existing theories or prior research, in all included studies. In building psychometric instruments, all reports attempted to understand human behaviour through specific emotions instead of following a more agnostic approach of gathering all possible emotions that may emerge from the studied population. In the surveyed instruments, there is no consensus on considering multiple components of emotions, which are affective, expressive behavioural, cognitive, motivational, and physical aspects.^64^ It has been further pointed out that using lists of emotion-words as done in some cases^40^ implies that single items are more vulnerable to random measurement errors and unknown biases in meaning and interpretation compared to multiple-item scales.^65,66^ Moreover, single-worded emotions might not accurately capture individual emotional experience as the interpretation by the respondent and the researcher may be ambiguous. On the other hand, the CCAS^15^ which has been the most used one in the literature, included only one emotion-related word (‘concern’) in the items, and derives them uniquely by how day-to-day experiences are impacted by climate anxiety. Therefore, there are methodological differences in measuring constructs with single item or multiple items. Employing the term “emotion” without qualification leads to misinterpretations and inconsistencies in both theoretical frameworks and research endeavours.^67^

Furthermore, all the validations of the included instruments reported a wide range of scales against which the new instrument was evaluated. This does not help comparison and consistency between instruments as it might imply attempting assessing a very wide range of severity of the same emotions/condition. Ideally, first the most comprehensive definition of eco-emotions should be established, and then to what extent these eco-emotions are related to people’s daily behaviours and attitudes regarding the environment should be tested. Using a bottom-up approach by starting with specific observations or data and working upward to develop a measurement tool would enable the creation of a multidimensional construct including behavioural, cognitive, and physiological components of each emotion, aiding the in-depth understanding of their distributions.

It is likely that in many of the populations (in particular among older generations) the prevalence of negative eco-emotions, including eco-anxiety, suggests a need for healing rather than viewing it as a condition necessitating fixing or curing.^20^ During our search process, we retrieved studies that used previously developed tools, such as state-trait anxiety scale to measure climate change anxiety^68,69^ and general guilt and shame scale to measure eco-guilt.^70^ Although participants were asked to report their emotions by taking climate change into consideration, these studies did not provide evidence of validity as to whether a specific anxiety for environmental incidents was measured. To obtain an accurate assessment of the relationship between climate change distress and other constructs (i.e. environmental concern and general anxiety), a valid measurement tool is needed. This will also allow us to define what the nosological entity of the condition is and to evaluate the impact of therapeutic responses for people whose anxiety is extreme.^15^ Moreover, an instrument developed for this specific purpose should be tailored not only to assess clinically relevant symptoms of anxiety, but mostly sub-clinical ones. For example, HEAS^17^ and CCAS^15^ contain items such as rumination or inability to perform daily activities, and complaints of close ones about these situations. Although those scales did not consider eco-anxiety as a pathological construct, it is possible that the severity level of the anxiety they measure is high.^39,71^ Importantly, all the emotions covered by the surveyed instruments are negative ones, making it difficult to assess the potential role of positive emotions such as hopefulness, empathy, humour, anticipated pride, gratitude, optimism, or feeling strong to do something though one’s contributions have been shown in taking an action for the environment.^72–74^ The importance of considering not only positive emotions but also other eco-emotions (e.g., hostility, disgust) has been emphasised.^72,75^ The need of thoroughly exploring a wide range of emotions without necessarily attributing them to a positive or negative quality is also motivated by the emerging idea that it might be important to focus more on anger, rather than anxiety.^76^ According to the current findings of general anger studies, there is a positive relationship between the level of anger and goal achievement in challenging tasks.^77^ It might be possible to speculate therefore that if we want to assess to what extent which emotion(s) trigger people to take action to reverse the environmental conditions or at least slow down the progress, as a first step, we must enable people to express all their feelings about changes in climate and environmental conditions. Followed by a preliminary study conducted to gather and cluster various emotions,^30^ a very recent instrument titled Inventory of Climate Emotions (ICE),^78^ published after the searches for the current review had been conducted, offers a relatively wider range of emotions related to climate change, including hopefulness and empowerment as positive emotional dimensions.

### Strengths and limitations of the current review

At least three main limitations to this study were identified. First, some instruments^78^ and validation studies^79,80^ published after the searches were run were not included. Second, despite the effort of capturing literature assessing all possible emotions, some studies might have been overlooked due to many different names of emotions used interchangeably in the literature. Third, qualitative studies were out of the scope of this study. However, they may have presented additional information about individual’s emotions associated with the climate crisis.

To ensure that emotions which might not have been included in the search string, such as fear, shame, and sadness were still captured by this review, the same search string was adapted to check if there was any tool measuring specifically eco-fear, eco-shame or eco-sadness and the search was run in the same databases, with no additional items identified.

### Implications and Future Research and Conclusions

Our findings reinforce the necessity of a bottom-up approach of the operational definitions of emotions and their environmental context. A comprehensive measure of eco-emotions is necessary to understand the diverse roles of each emotion in different outcomes (e.g., mental health, taking individual or political action, life decision-making processes) and in different populations (e.g., young vs. old generations, Global South/North, people exposed to direct or indirect effects of climate crises) or to evaluate the efficiency of educational programs that provide information on climate change, where individuals actively engage in generating individual and societal policies, and also strive to adopt and promote behaviours that mitigate climate change. The psychometric instrument should focus on sub-clinical traits to increase the generalizability of the findings. The tools should be adaptable into different cultures and potential cross-cultural variations should be tested.

The newly developed ICE^78^ fulfilled these requirements, and took into account, most of the methodological limitations of previous studies. By conducting a preliminary study, providing clear definitions for each emotion, involving behavioural, cognitive, and affective components of emotions in the items, taking not only the negative but also the positive emotions into account, and testing various types of validity of the tool, it represents a comprehensive measure worth testing in different samples and languages. However, considering that the levels and forms of exposure to the climate crisis differ day to day in different geographies and that precautions and policies vary between countries, the diversity of emotions and the ways they are expressed are worth examining delicately.

## Data Availability

All data produced in the present work are contained in the manuscript.

## Acknowledgement

The first author was funded by the Scientific and Technological Research Council of Turkey (TÜBİTAK) to carry out postdoc research at the time this study was conducted (Grant Number: 1059B192202875). The funding source had no specific role in study design, in the writing of the report; or in the decision to submit the paper for publication.

---

**Box 1:**
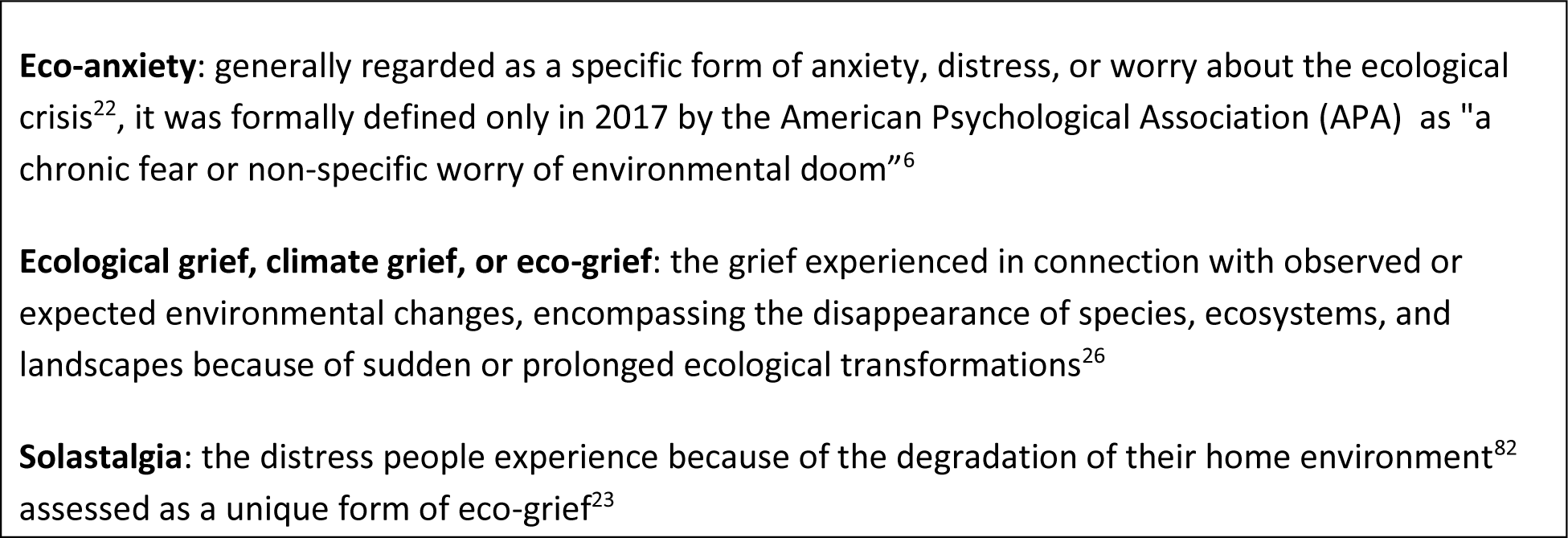
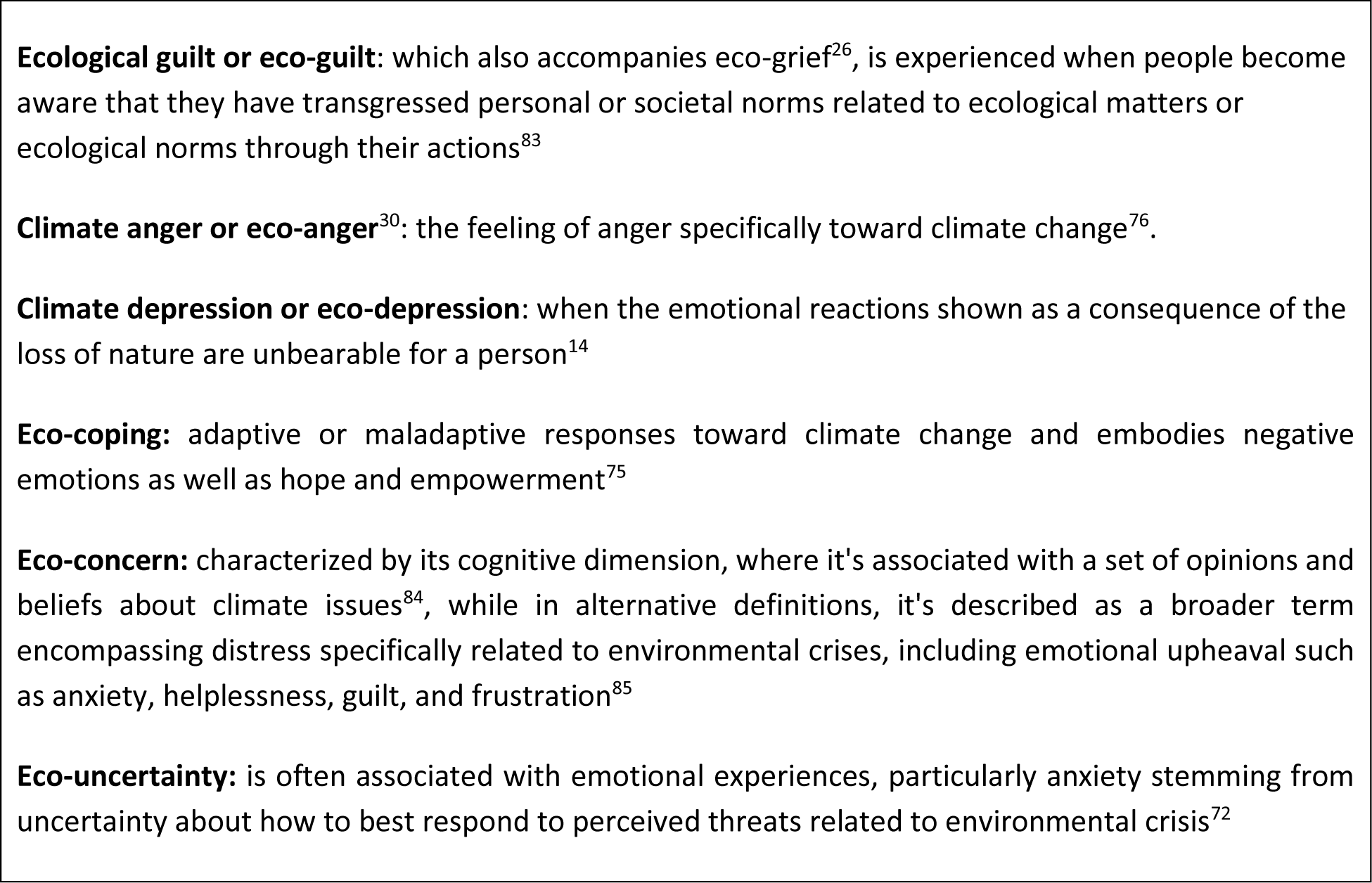
Definition of the different eco-emotions.

