## Supplementary Material 1 for "Measuring Planetary Eco-Emotions: A Systematic Review of Currently Available Instruments and Their Psychometric Properties"

*Validation of Climate Change Anxiety Scale (CCAS) in additional populations*

The validation of 22-item version of CCAS was tested with samples from the US[1], France[2], and Australia[3], with one group concluding that it was statistically more appropriate to use the 13-item version[2]. Other studies directly tested the 13-item version with different factorial solutions in samples from Germany[4], Italy[5], France[2], Philippines[6], Poland[7], Korea[8], US, China, India, and Japan[9]. The four-item version of the CCAS was tested with Canadian adolescents[10], and the six-item version was tested in a US sample[11]. These studies suggested different factorial structures for the scale. While Hogg et al.[12] supported the two-factor structure for the 13-item version of it, as a result of the factor analyses and high correlation between the two subscales of the 13-item version of CCAS, three studies suggested not to treat the two subscales (i.e., cognitive-emotional impairment and functional impairment) as separate constructs but to use a one-factor structure[4,5,11]. Although Cruz and High[11] conducted their studies in the same country as the original study, they did not replicate the same factor structure as Clayton and Karazsia[1]. Four of the studies conducted in different countries supported the two-factor structure[2,6,8,9], whereas one study suggested three factors[7].

*Test of the factor structures*

To test the factor structure and the model fit to the data, exploratory factor analysis (EFA) and confirmatory factor analysis (CFA) were conducted for six scales in scale development studies[1,12–14], while three scale development studies carried out only EFA[15–17] and one did not report any factor analysis result[18]. Three scale development studies[12,16,17] and two scale validation studies[8,19] reported performing EFA using Principal Component Analysis (PCA) method. Among the scale validation studies, five tested both EFA and CFA[5,7,8,10,19], and six conducted only CFA[2–4,6,9,11], while one did not report any factor analysis result[20]. Among those conducted both EFA and CFA, four studies used different samples to conduct the analyses[1,10,12,13], while the other five conducted EFA and CFA in the same sample. Multigroup CFA was conducted by two studies[3,9]. For those conducted CFA, the model fit indices were indicated on Table 2. The Tucker–Lewis index (TLI), the comparative fit index (CFI), the root mean square error of approximation (RMSEA), and the standardized root mean squared residual (SRMR) were used to evaluate the measurement models. Acceptable values for CFI and TLI are typically above .90 or .95, and for SRMR below .08[21]. RMSEA values below .05 indicate a good fit[22] and below .08 indicate appropriate fit[21]. To these criteria, one of the scale validation studies showed poor CFI[5]. Except for four studies that did not report the TLI value[5,11,14,19], all met the TLI criteria. Eight studies did not report SRMR, whereas the other nine provided the rule of thumb[2,4,6–12]. To the RMSEA criteria, one study showed a good fit[14] and seven studies indicated an appropriate fit (eco-anxiety, eco-guilt, and ecological grief[1,2,4,6,7,12,13] while one did not report RMSEA[11].

*Test of validity*

The studies testing the convergent or discriminant validity of the instruments employed various types of variables. The most frequently used variables were anxiety (generalized, trait, state, or future anxiety) used in ten studies and depression used in nine studies. Other emotional/behavioural/cognitive dimensions were also used, such as stress, environmental identity, and pro-environmental behaviours. Two studies compared the groups of people who were exposed and those who were not exposed to environmental damage. One study involving participants from more than one country provided evidence for the cross-cultural validity of CCAS[9]. The variables used to test the validity were indicated in Table 2. The results of analyses conducted to test factorial structures and validity of each scale were presented in Table 3.

**Table 3.** Factor analyses and validity testing results from the studies testing the validity of the scales

| Name of the scale | Source | Country / Language | Number of items | Number and names of factors (number of items per factor) / analysis type / sample size | Variables used to gather validity evidence on test relationships and the results of analyses for each (sub)scale |
|---|---|---|---|---|---|
| Environmental Worry Scale (EWS)<br>(Scale development) | Bowler & Schwarzer (1991) | USA / English | 17 and 8 (for short version) | one-factor (environmental worry)<br><br>EFA or CFA not conducted<br>n = 547<br>(pretested in a sample of 250 undergrads) | EWS17<br>tension: 0.40<br>depression: 0.35<br>anger: 0.35<br>vigor: -0.35<br>fatigue: 0.35<br>confusion: 0.39<br>anxiety: 0.35<br>exposure: 0.37<br>EWS8<br>tension: 0.48<br>depression: 0.44<br>anger: 0.42<br>vigor: -0.41<br>fatigue: 0.42<br>confusion: 0.48<br>anxiety: 0.44<br>exposure: 0.46<br>p-values: NR |
| Environmental Distress Scale (Solastalgia subscale) *<br>(Scale development) | Higginbotham et al. (2007) | Australia / English | 9 | one-factor for solastalgia (9)<br><br>EFA using PCA one-factor solution for solastalgia subscale<br>n =203 | Comparison of non-exposed vs. exposed groups<br>Non-exposed (M = 20.8 (6.3) reported less solastalgia exposed (M = 28.3 (7.2), t= -7.71; $p < 0.001$ |
| Climate Change Distress Scale<br>(Scale development) | Searle & Gow (2010) | Australia / English | 12 | two-factor<br>(climate change anxiety (9), climate change hopelessness (3))<br>EFA using PCA with oblique rotation<br>n = 275 | Hierarchical regression analyses for climate change anxiety and climate change hopelessness, respectively<br>environmental beliefs:<br>$\beta = 0.33$; $\beta = 0.22$, $p < 0.001$<br>future anxiety:<br>$\beta = 0.38$; $\beta = 0.53$, $p < 0.001$ |

| | | | | | |
|---|---|---|---|---|---|
| | | | | | intolerance of uncertainty:<br>$\beta$ -0.10; $\beta$ = -0.16, p > .05<br>religiosity:<br>$\beta$ = -0.01; $\beta$ = -0.05, p > .05 |
| Climate Change Anxiety Scale (CCAS)<br>(Scale development) | Clayton & Karazsia (2020) | USA / English | 22 | four-factor<br>(cognitive-emotional impairment (8), behavioural engagement (6), experience (3), functional impairment (5))<br>EFA using PAF with oblimin rotation suggesting four factor solution<br>n=197<br><br>CFA<br>CFI = 0.93<br>TLI = 0.92<br>SRMR = NR<br>RMSEA = 0.07, CI 90% [0.06 − 0.08]<br>n=199 | Correlational analyses to test concurrent and discriminant validity of cognitive-emotional impairment, behavioural engagement, experience, and functional impairment, respectively<br>environmental identity:<br>r = 0.22, r = 0.53, r = 0.46, r = 0.17, p < .05<br>negative emotionality:<br>r = 0.52, r = 0.23, r = 0.37, r = 0.45, p < .05<br>depression/anxiety:<br>r = 0.54, p < .05; r = 0.01, p > .05; r = 0.16, p < .05; r = 0.47, p < .05 |
| CCAS<br>(Scale validation) | Innocenti et al. (2021) | Italy / Italian | 13 | two-factor used (cognitive-emotional impairment (8), functional impairment (5))<br>EFA recommended one-factor<br>n = 150<br>CFA to test two-factor model<br>CFI = 0.754<br>TLI = NR<br>SRMR = NR<br>RMSEA = 0.126 (0.106−0.145, p < 0.001) | ANCOVA to test convergent validity of cognitive-emotional impairment and functional impairment, respectively<br>general anxiety disorder:<br>B = 0.898, p = 0.001; B = 1.349, p = 0.006<br>distress:<br>B = 0.437, p = 0.013; B = 0.599, p = 0.059<br>depression:<br>B = 0.297, p= 0.011; B = 0.367, p = 0.081<br>anxiety:<br>B = 0.141, p =0.036; B = 0.232, p = 0.054<br>new social paradigm (New Ecological Paradigm (NEP) subscale):<br>B = 0.292, p = 0.001; B = 0.173, p = 0.263<br>ANCOVA to test concurrent validity<br>self-efficacy: |

| | | | | | |
|---|---|---|---|---|---|
| | | | | | B = -0.381, p < 0.001; B = -0.454, p = 0.014<br>dominant paradigm (NEP subscale): subscale:<br>B = -0.286, p = 0.008; B = -0.248, p = 0.051<br>pro-environmental behaviours:<br>B = 1.452, p < 0.001; B = 1.933, p < 0.001 |
| CCAS<br>(Scale validation) | Wullenkord et al. (2021) | Germany / German | 13 | one-factor<br>(climate change anxiety)<br>CFA<br>Robust CFI= 0.92<br>Robust TLI= 0.90<br>SRMR= 0.051<br>Robust RMSEA= 0.85 90% CI [0.074, 0.095]<br>n = 1011 | pro-environmental intentions:<br>β = 0.43, p < 0.001<br>avoidance:<br>β = 0.21, p < 0.001<br>denial of personal outcome severity of climate change:<br>β = 0.08, p < 0.041<br>human dominance over nature:<br>β = 0.11, p < 0.001<br>general anxiety and depression:<br>β = 0.10, p = 0.004<br>competence frustration:<br>β = 0.09, p = 0.026<br>Right-wing political orientation:<br>β = 0.06, p = 0.049<br>denial of guilt:<br>β = − 0.26, p < 0.001 |
| CCAS<br>(Scale validation) | Cruz & High (2022) | USA / English | 13 (does not fit)<br>(11-item restructured version best fit) | one-factor (climate anxiety)<br><br>CFA<br>CFI = .99<br>TLI = NR<br>SRMR .02<br>RMSEA = NR<br>n = 513 | Correlational analyses to test discriminant validity of climate anxiety:<br>depression: r = .36<br>trait anxiety r = .32<br>state anxiety: r = .41<br>p-values not reported |
| CCAS<br>(Scale validation) | Larionow (2022) | Poland / Polish | 13 | three-factor (functional impairment (5),<br>intrusive symptoms (4), reflections on climate anxiety (4)<br><br>EFA with oblimin rotation with 3-factor solution | Correlational analyses to test validity of functional impairment, intrusive symptoms, and reflections on climate anxiety, respectively (n ranges from 64 to 137)<br>Intrusive symptoms:<br>experience of climate change:<br>r = 0.45, p < 0.001; r = 0.26, p < 0.05; r = 0.50, p < 0.001<br>behavioural engagement: |

| | | | | | |
|---|---|---|---|---|---|
| | | | | CFA<br>CFI = 0.968<br>TLI = 0.959<br>SRMR = 0.040<br>RMSEA = 0.062 (0.048–0.07)<br>n = 603 | r = 0.40, p < 0.001; r = 0.34, p < 0.01; r = 0.43, p < 0.001<br>environmental identity:<br>r= 0.49, p < 0.001; r = 0.28, p < 0.01; r = 0.35, p < 0.001<br>biospheric concerns:<br>r = 0.46, p < 0.001; r = 0.28, p < 0.05; r = 0.43, p < 0.001<br>altruistic concerns:<br>r = 0.34, p < 0.001; r = 0.24, p > 0.05; r = 0.22, , p > 0.05<br>egoistic concerns:<br>r = 0.37, p < 0.001; r = 0.24, p > 0.05; r = 0.34, p < 0.01<br>climate change denial:<br>r = -0.57; r = -0.50; r = -0.55, p < 0.001<br>anxiety symptoms:<br>r = 0.10; r = 0.11; r = 0.03, p > 0.05<br>depressive symptoms:<br>r = 0.27, p < 0.01; r = 0.20, p < 0.05; r = 0.14, p > 0.05<br>anxiety-depressive symptoms:<br>r = 0.20, p < 0.05; r = 0.17, p > 0.05; r = 0.10, p > 0.05<br>sense of safety:<br>r = -0.35, p < 0.001; r = 0.20, p < 0.05; r = -0.29, p < 0.01<br>self-blame:<br>r = 0.03; r = 0.17; r = 0.14, p > 0.05<br>acceptance:<br>r = 0.04; r = 0.05; r = -0.18, p > 0.05<br>rumination:<br>r = -0.06; r = -0.05; r = -0.18, p > 0.05<br>positive refocusing:<br>r = 0.11; r = 0.15; r = 0.07, p > 0.05<br>refocus on planning:<br>r = 0.05; r = 0.10; r = 0.18, p > 0.05<br>positive reappraisal:<br>r = 0.15; r = 0.14; r = 0.17, p > 0.05<br>putting into perspective:<br>r = 0.17; r = 0.19; r = 0.03, p > 0.05<br>catastrophizing:<br>r = 0.05; r = 0.10; r = -0.20, p > 0.05<br>blaming others:<br>r = 0.03; r = 0.05; r = 0.09, p > 0.05 |

| | | | | | |
|---|---|---|---|---|---|
| CCAS<br>(Scale validation) | Mouguiama-Daouda et al. (2022) | France / French | 13 | two-factor<br>(cognitive-emotional impairment (8), functional impairment (5))<br><br>CFA<br>CFI = 0.92<br>TLI = 0.91<br>SRMR = 0.05<br>RMSEA = 0.07 (0.061–0.093)<br>n = 305, n = 905 | Correlational analyses for cognitive-emotional impairment and functional impairment, respectively:<br>depression:<br>$r = 0.28$; $r = 0.27$, $p < .05$<br>general anxiety disorder:<br>$r = 0.05$; $r = -0.03$, $p > .05$<br>Environmental identity:<br>$r = 0.34$; $r = 0.29$ $p < .05$ |
| CCAS<br>(Scale validation) | Simon et al. (2022) | Philippines / English | 13 | two-factor<br>(cognitive-emotional impairment (8), functional impairment(5))<br><br>CFA<br>CFI = 0.972<br>TLI= 0.963<br>SRMR = 0.041<br>RMSEA = 0.060<br>n = 452 | Convergent validity based on computations of composite reliability (CR) and average variance extracted (AVE)<br>Cognitive Emotional CR: 0.89<br>Cognitive Emotional AVE: 0.51<br>Functional CR: 0.85<br>Functional AVE: 0.53<br>Discriminant validity based on maximum shared variance (MSV)<br>Cognitive Emotional MSV: 0.74<br>Functional MSV: 0.74<br>The two subscales did not meet the criteria for discriminant validity |
| CCAS<br>(Scale validation) | Jang et al. (2023) | Korea / Korean | 13 | two-factor<br>(cognitive-emotional impairment (8), functional impairment (5))<br><br>EFA using PCA with varimax orthogonal rotation<br>n = randomly selected 350 out of 459<br><br>CFA<br>CFI = 0.94<br>TLI = 0.92<br>SRMR = 0.05<br>RMSEA = 0.09 | Standardized<br>factor loading (β), average variance extracted (AVE), and<br>composite reliability (CR) were used for convergent validity<br>of each item, and the correlational analyses and AVE value<br>were used for discriminant validity.<br>β was 0.64 to 0.83 (> 0.50)<br>AVE ranged from 0.50 to 0.58 (> 0.50)<br>CR was 0.86–0.91 (> .70)<br>The $r^2$ of Factor 1 items was 0.46–0.62 (smaller than the AVE of 0.75 for Factor 1), and the $r^2$ of Factor 2 items |

| | | | | | |
|---|---|---|---|---|---|
| | | | | n = 459 | was 0.38–0.56 (smaller than the AVE of 0.58 for Factor 2). |
| CCAS<br>(Scale validation) | Tam et al.<br>(2023) | China, India, Japan, and USA / English, Chinese, Japanese | 13 | two-factor<br>(cognitive-emotional impairment (8), functional impairment (5))<br><br>CFA two-factor<br>**China**<br>Robust RMSEA = 0.084<br>Robust CFI = 0.954<br>SRMR = 0.033<br>**India**<br>Robust RMSEA = 0.087<br>Robust CFI = 0.931<br>SRMR = 0.043<br>**Japan**<br>Robust RMSEA = 0.134<br>Robust CFI = 0.900<br>SRMR = 0.050<br>**US**<br>Robust RMSEA = 0.115<br>Robust CFI = 0.936<br>SRMR = 0.035<br>**Multigroup CFA**<br>Configural Invariance<br>RMSEA (ΔRMSEA) = 0.102 (-)<br>CFI (ΔCFI) = 0.936 (-)<br>SRMR (ΔSRMR) = 0.037 (-)<br>Metric invariance<br>RMSEA (ΔRMSEA) = 0.099 (-.003)<br>CFI (ΔCFI) = 0.933 (-.003)<br>SRMR (ΔSRMR) = 0.054 (.017)<br>Scalar invariance<br>RMSEA (ΔRMSEA) = 0.103 (.004) | Correlational analyses for cognitive-emotional impairment and functional impairment, respectively per country<br>**China**<br>climate change beliefs<br>climate change belief in happening:<br>r= -.043, p > .05; r = -.106, p < .001<br>climate change belief in scientific consensus:<br>r = -.033, p > .05; r = -.080, p < .001<br>worry:<br>r = .333; r = .224, p < .001<br>perceived harm to self:<br>r = .0311; r = .209, p < .001<br>perceived harm to country:<br>r = .222; r =.109, p < .001<br>climate action:<br>resource conservation:<br>r = .147, p < .001; r = .058, p > .05<br>sustainable diet:<br>r = .396; r = .406, p < .001<br>climate activism:<br>r = .507; r= .433, p < .001<br>support for climate policy:<br>r = .325; r = .300, p < .001<br>**India**<br>climate change beliefs<br>belief in happening:<br>r = .028; r = -.031, p > .05<br>belief in scientific consensus r = .113, p < .001; r = .044, p > .05<br>worry:<br>= r =.333; r =.235, p < .001 |

| | | | | | |
|---|---|---|---|---|---|
| | | | | CFI (ΔCFI) = 0.918 (-.015)<br>SRMR (ΔSRMR) = 0.064 (.010)<br><br>N = 4000 (1000 from each country) | perceived harm to self:<br>r = .319; r = .244, p < .001<br>perceived harm to country:<br>r = .248; r =.174, p < .001<br>climate action<br>resource conservation:<br>r = .218, p > .05; r = .137, p < .001<br>sustainable diet:<br>r = .349; r = .317, p < .001<br>climate activism:<br>r =.444; r = .353, p < .001<br>support for climate policy:<br>r = .182, p < .001; r = .061, p > .05<br>**Japan**<br>climate change beliefs<br>belief in happening:<br>r = .119, p < .001; r = .093, p < .01<br>belief in scientific consensus:<br>r = .127, p < .001; r = .105, p < .01<br>worry:<br>r = .283; r =.200, p < .001<br>perceived harm to self:<br>r = .261; r = .202, p < .001<br>perceived harm to country:<br>r = .225; r = .142, p < .001<br>climate action<br>resource conservation:<br>r = .208; r = .131, p < .001<br>sustainable diet:<br>r = .419; r = .352, p < .001<br>climate activism:<br>r = .551; r = .489, p < .001<br>support for climate policy:<br>r = .278; r = .217, p < .001<br>**USA**<br>climate change beliefs<br>belief in happening: |

| | | | | | |
|---|---|---|---|---|---|
| | | | | | r = .166; r = .123, p < .001<br>belief in scientific consensus<br>r = .084, p < .05; r = .045<br>worry:<br>r = .330; r =.246, p < .001<br>perceived harm to self:<br>r = .409; r = .334, p < .001<br>perceived harm to country:<br>r = .308; r = .234, p < .001<br>climate action<br>resource conservation: r = .156, p < .001; r = .102, p < .01<br>sustainable diet:<br>r = .476; r = .433, p < .001<br>climate activism:<br>r = .542; r = .463, p < .001<br>support for climate policy:<br>r = .282; r = .211, p < .001 |
| CCAS<br>(Scale validation) | Wu et al.<br>(2023) | Canada /<br>English | 4 | one-factor<br>(climate anxiety)<br><br>EFA<br>n = 1144<br><br>CFA<br>Robust CFI = 0.994<br>TLI = NR<br>SRMR = 0.006<br>RMSEA = 0.178 (90% CI [0.103, 0.268])<br>n = 1162 | Correlational analyses to test convergent validity of climate anxiety<br>general anxiety:<br>0.17, p = 0.0001<br>depression:<br>r = 0.14, p = 0.0001<br>climate concern:<br>r = 0.24, p = 0.0001<br>positive mental health:<br>r = -0.09, p = 0.0001<br>life satisfaction:<br>r = 0.025, p > .05<br>Correlational analysis to test discriminant validity of climate anxiety<br>Self-concept:<br>r = -0.05, p = 0.04 |
| Hogg Eco-Anxiety Scale<br>(HEAS) | Hogg et al.<br>(2021) | New Zealand<br>/ English | 13 | four-factor (affective symptoms (4), rumination (3), behavioural | Correlational analyses to test concurrent and discriminant validity of affective symptoms, rumination, |

| | | | | | |
|---|---|---|---|---|---|
| (Scale development) | | | | symptoms (3), personal impact anxiety (3))<br><br>EFA using PCA with oblimin rotation<br>n = 343<br><br>CFA<br>CFI = 0.96<br>TLI = 0.95<br>SRMR = 0.07<br>RMSEA = 0.08 (90% CI [0.07 – 0.10])<br>n = 342 | behavioural symptoms, personal impact anxiety, respectively<br>stress:<br>0.42; 0.22; 0.30; 0.27; p < 0.001<br>anxiety:<br>0.46; 0.22; 0.31; 0.28, p<0.001<br>depression:<br>0.37, p < 0.001; 0.15, p < 0.01; 0.35, p < 0.001; 0.21, p < 0.001<br>emotional reactivity:<br>0.37, p < 0.001; 0.13, p < 0.05; 0.23, p < 0.001; 0.20, p < 0.001<br>credibility of science:<br>-0.05; 0.01; 0.07; 0.05, p > .05<br>climate change belief:<br>0.06, p > .05; 0.14, p < 0.05; 0.01, p > .05; 0.23, p < 0.001 |
| HEAS<br>(Scale validation) | Uzun et al., 2022 | Türkiye/Turkish | 13 | four-factor<br>(affective symptoms (4), rumination (3), behavioural symptoms (3), personal impact anxiety (3))<br><br>EFA by using PCA<br>n = 698<br>CFA<br>CFI = 0.97<br>TLI = NR<br>SRMR = NR<br>RSMEA = 0.06 | average variance extracted (AVE), and composite reliability (CR) for affective symptoms, rumination, behavioural symptoms, personal impact anxiety, respectively<br><br>AVE: 0.59; 0.63; 0.62; 0.64<br>CR: 0.81; 0.83; 0.86; 0.84 |
| HEAS<br>(Scale validation) | Pavani et al., 2023 | France / French | 13 | one-factor<br>(eco-anxiety)<br><br>EFA or CFA not conducted<br>n = 200 | pro-environmental behaviours on eco-anxiety at time 1:<br>B = 0.153, p = 0.004 |

| | | | | | |
|---|---|---|---|---|---|
| CCAS and HEAS<br>(Scale validation) | Hogg et al., 2023 | Australia / English | 22 for CCAS & 13 for HEAS | four-factor<br>(affective symptoms (4), rumination (3), behavioural symptoms (3), personal impact anxiety (3)) for HEAS and two-factor (cognitive-emotional impairment (8), functional impairment (5)) for CCAS with 13-item<br>Multigroup CFA<br>HEAS:<br>Gender:<br>ΔCFI ≤ 0.005<br>ΔRMSEA ≤ 0.010<br>ΔSRMR ≤ 0.025 for metric and ≤0.005 for scalar models<br>Age:<br>ΔCFI ≤ 0.010 ΔRMSEA ≤0.015<br>ΔSRMR: ≤ 0.030 for metric and ≤ 0.010 for scalar<br><br>CCAS<br>Gender:<br>The metric and scalar models (CFI < 0.90, RMSEA > 0.10)<br>Age:<br>Configural, metric and scalar (CFI < 0.90, RMSEA > 0.10)<br>n = 530 | Correlational analyses for affective symptoms, rumination, behavioural symptoms, personal impact anxiety, cognitive-emotional impairment, and functional impairment, respectively<br>risk perception:<br>r = .41; r = .45; r = .30; r = .43, r = .38; r = .39, p < .001<br>direct experience:<br>r =.30; r =.37; r =.30; r =.26; r =.35; r =.33, p < .001<br>see information:<br>r =.20; r =.25; r =.16; r =.18; r =.17; r =.21, p < .001<br>seek information:<br>r =.26; r =.36; r =.23; r =.25; r =.32; r =.34, p < .001<br>avoid information:<br>r =.18, p < .001; r =.10, p < .01, r =.15, p < .001; r =.15, p < .001 r =.14, p < .001; r =.16, p < .001 |
| Eco-Anxiety Questionnaire<br>(Scale development) | Agoston et al., 2022 | Hungary / Hungarian | 22 | two-factor (habitual ecological worry (13), negative consequences of anxiety (9))<br><br>EFA with WLSMV estimation and geomin rotation<br>n = 1152 out of 4608<br><br>CFA with WLSMV estimation | Correlational analyses for habitual ecological worry and negative consequences of anxiety<br>Pro-environmental behaviours<br>Sorting trash into the recycling:<br>r = 0.218; r = 0.120, p < 0.01<br>Composting or reusing household food garbage:<br>r = 0.131; r = 0.162, p < 0.01<br>Using reusable bags:<br>r = 0.175; r = 0.111, p < 0.01 |

| | | | | | |
|---|---|---|---|---|---|
| | | | | CFI = 0.972<br>TLI = 0.969<br>SRMR = NR<br>RMSEA = 0.056, 90%CI [0.052 - 0.059]<br>n = 1152 out of 4608 | Eating meat:<br>r = 0.203; r = 0.231, p < 0.01<br>Eating dairy products or egg:<br>r = 0.103; r = 0.150, p < 0.01<br>Walking, cycling, or taking transportation instead of using a car:<br>r = 0.143; r = 0.112, p < 0.01<br>Saving energy:<br>r = 0.164; r = 0.141, p < 0.01<br>Conserving water:<br>r = 0.230; r = 0.152, p < 0.01<br>Using second-hand clothes:<br>r = 0.174; r = 0.222, p < 0.01 |
| Eco-Guilt Questionnaire (Scale development) | Agoston et al., 2022 | Hungary / Hungarian | 11 | one-factor<br>(eco-guilt)<br>EFA with WLSMV estimation and geomin rotation<br>n = 1152 out 4608<br>CFA with WLSMV estimation<br>CFI = 0.986<br>TLI = 0.977<br>SRMR = NR<br>RMSEA = 0.077, 90%CI [0.069, 0.084]<br>n = 1152 out of 4608 | Pro-environmental behaviours<br>Sorting trash into the recycling:<br>r = 0.124<br>Composting or reusing household food garbage:<br>r = 0.051<br>Using reusable bags:<br>r = 0.094<br>Eating meat:<br>r = 0.138<br>Eating dairy products or egg:<br>r = 0.053<br>Walking, cycling, or taking transportation instead of using a car:<br>r = 0.079<br>Saving energy:<br>r = 0.056<br>Conserving water:<br>r = 0.077<br>Using second-hand clothes:<br>r = 0.120<br>All are significant at p < 0.01 |

| | | | | | |
|---|---|---|---|---|---|
| Ecological Grief Questionnaire (Scale development) | Agoston et al., 2022 | Hungary / Hungarian | 6 | one-factor (ecological grief)<br><br>EFA with WLSMV estimation and geomin rotation<br>n = 1152 out of 4608<br>CFA with WLSMV estimation<br>CFI = 0.986<br>TLI = 0.977<br>SRMR = NR<br>RMSEA = 0.064, 90%CI[0.047, 0.081]<br>n = 1152 out of 4608 | Correlational analyses for ecological grief<br>pro-environmental behaviours<br>sorting trash into the recycling:<br>r = 0.168<br>composting or reusing household food garbage:<br>r = 0.173<br>using reusable bags:<br>r = 0.151<br>eating meat:<br>r = 0.182<br>eating dairy products or egg:<br>r = 0.091<br>walking, cycling, or taking transportation instead of using a car:<br>r = 0.088<br>saving energy:<br>r = 0.170<br>conserving water:<br>r = 0.227<br>using second-hand clothes:<br>r = 0.187<br>All are significant at $p < 0.01$ |
| Climate Change Worry Scale (Scale development) | Stewart, 2021 | USA / English | 10 | one-factor (climate change worry)<br><br>EFA<br>n = 600<br><br>CFA<br>CFI = 0.99<br>TLI = NR<br>SRMR = NR<br>RMSEA = 0.043 | Correlational analyses to test convergent and divergent validity of climate change worry<br>political orientation:<br>r = -0.43, $p < 0.0001$<br>fear of Weather:<br>r = 0.30, $p < 0.0001$<br>storm fear:<br>r = 0.30, $p < 0.0001$<br>stress:<br>r = 0.31, $p < 0.0001$<br>anxiety:<br>r = 0.29, $p < 0.0001$<br>depression:<br>r = 0.30, $p < 0.0001$<br>worry: |

| | | | | | |
|---|---|---|---|---|---|
| | | | | | r = 0.17, p < 0.05<br>n = 353 (study 3) |
| Scale of Solastalgia<br>(Scale development) | Caceres et al., 2022 | Chile / no info. | 10 | two-factor (solace (7), algia (3))<br>EFA by using PFA with oblimin rotation<br>n = 223 | Correlational analyses for solace and algia post-traumatic stress disorder:<br>r = 0.150, p < 0.05; r = 0.359, p < 0.01 |

*Note.* NR: Not reported, PCA: Principal Component Analysis, PFA: Principal Factor Analysis.

* The solastalgia subscale of the Environmental Distress Scale was involved in this review based on the scope of the current review.

1 Clayton S, Karazsia BT. Development and validation of a measure of climate change anxiety. *Journal of Environmental Psychology* 2020; **69**: 101434.

2 Mouguiama-Daouda C, Blanchard MA, Coussement C, Heeren A. On the Measurement of Climate Change Anxiety: French Validation of the Climate Anxiety Scale. *PSYCHOL BELG* 2022; **62**: 123.

3 Hogg TL, Stanley SK, O'Brien LV. Synthesising psychometric evidence for the Climate Anxiety Scale and Hogg Eco-Anxiety Scale. *Journal of Environmental Psychology* 2023; **88**: 102003.

4 Wullenkord MC, Tröger J, Hamann KRS, Loy LS, Reese G. Anxiety and climate change: a validation of the Climate Anxiety Scale in a German-speaking quota sample and an investigation of psychological correlates. *Climatic Change* 2021; **168**: 20.

5 Innocenti M, Santarelli G, Faggi V, *et al.* Psychometric properties of the Italian version of the Climate Change Anxiety Scale. *The Journal of Climate Change and Health* 2021; **3**: 100080.

6 Simon PD, Pakingan KA, Aruta JJBR. Measurement of climate change anxiety and its mediating effect between experience of climate change and mitigation actions of Filipino youth. *Educational and Developmental Psychologist* 2022; **39**: 17–27.

7 Larionow P, Sołtys M, Izdebski P, *et al.* Climate Change Anxiety Assessment: The Psychometric Properties of the Polish Version of the Climate Anxiety Scale. *Front Psychol* 2022; **13**: 870392.

8 Jang SJ, Chung SJ, Lee H. Validation of the Climate Change Anxiety Scale for Korean Adults. *Perspectives in Psychiatric Care* 2023; **2023**: 1–8.

9 Tam K-P, Chan H-W, Clayton S. Climate change anxiety in China, India, Japan, and the United States. *Journal of Environmental Psychology* 2023; **87**: 101991.

10 Wu J, Long D, Hafez N, Maloney J, Lim Y, Samji H. Development and validation of a youth climate anxiety scale for the Youth Development Instrument survey. *Int J Mental Health Nurs* 2023; : inm.13201.

11 Cruz SM, High AC. Psychometric properties of the climate change anxiety scale. *Journal of Environmental Psychology* 2022; **84**: 101905.

12 Hogg TL, Stanley SK, O'Brien LV, Wilson MS, Watsford CR. The Hogg Eco-Anxiety Scale: Development and validation of a multidimensional scale. *Global Environmental Change* 2021; **71**: 102391.

13 Ágoston C, Urbán R, Nagy B, *et al.* The psychological consequences of the ecological crisis: Three new questionnaires to assess eco-anxiety, eco-guilt, and ecological grief. *Climate Risk Management* 2022; **37**: 100441.

14 Stewart AE. Psychometric Properties of the Climate Change Worry Scale. *IJERPH* 2021; **18**: 494.

15 Caceres C, Leiva-Bianchi M, Serrano C, Ormazábal Y, Mena C, Cantillana JC. What Is Solastalgia and How Is It Measured? SOS, a Validated Scale in Population Exposed to Drought and Forest Fires. *IJERPH* 2022; **19**: 13682.

16 Higginbotham N, Connor L, Albrecht G, Freeman S, Agho K. Validation of an Environmental Distress Scale. *EcoHealth* 2007; **3**: 245–54.

17 Searle K, Gow K. Do concerns about climate change lead to distress? *International Journal of Climate Change Strategies and Management* 2010; **2**: 362–79.

18 Bowler RM, Schwarzer R. Environmental anxiety: Assessing emotional distress and concerns after toxin exposure. *Anxiety Research* 1991; **4**: 167–80.

19 Uzun K, Ozturk AF, Karaman M, *et al.* Adaptation of the Eco-Anxiety Scale to Turkish: A Validity and Reliability Study. *Arch Health Sci Res* 2022; **9**: 110–5.

20 Pavani J-B, Nicolas L, Bonetto E. Eco-Anxiety motivates pro-environmental behaviors: a Two-Wave Longitudinal Study. *Motiv Emot* 2023; **47**: 1062–74.

21 Hu L, Bentler PM. Cutoff criteria for fit indexes in covariance structure analysis: Conventional criteria versus new alternatives. *Structural Equation Modeling: A Multidisciplinary Journal* 1999; **6**: 1–55.

22 Browne M, Cudeck R. Alternative ways of assessing model fit. In: Testing structural equation models. Sage, 1993: 136–62.
