## Supplementary Material 2 for "Measuring Planetary Eco-Emotions: A Systematic Review of Currently Available Instruments and Their Psychometric Properties"

COSMIN Study Design and Risk of Bias (RoB) evaluation tables are presented below.

| Name of the scale | Source | Measurement properties | Content validity | Structural validity | Internal consistency | Measurement invariance | Measurement error and reliability | Criterion validity | Hypotheses testing for construct validity | Responsiveness | Translation process | Overall Quality Rating |
|---|---|---|---|---|---|---|---|---|---|---|---|---|
| Environmental Worry Scale | Bowler & Schwarzer, 1991 | 30/30 | 5/ 27 | 0/12 | 3/12 | NA | 0/24 | NA | 32/36 | 52/54 | NA | Doubtful |
| Environmental Distress Scale | Higginbotham et al., 2007 | 30/30 | 10/27 | 8/12 | 10/12 | NA | 22/24 | NA | 15/15 | 24/24 | NA | Adequate |
| Climate Change Distress Scale | Searle & Gow, 2010 | 27/30 | 3/27 | 10/12 | 12/12 | NA | 0/24 | NA | 21/21 | 30/30 | NA | Adequate |
| Climate Change Anxiety Scale (CCAS) | Clayton & Karazsia, 2022 | 28/30 | 0/27 | 9/12 | 10/12 | NA | 0/24 | NA | 14/21 | 17/30 | NA | Adequate |
| Hogg Eco-Anxiety Scale (HEAS) | Hogg et al, 2021 | 30/30 | 10/27 | 10/12 | 10/12 | NA | 22/24 | NA | 19/21 | 22/24 | NA | Good |
| Eco-Anxiety, Eco-Guilt,and Ecological Grief Questionnaires* | Ágoston et al., 2022 | 28/30 | 23/27 | 10/12 | 10/12 | NA | 0/24 | NA | 21/21 | 30/30 | NA | Adequate |
| Climate Change Worry Scale | Stewart, 2021 | 27/30 | 0/27 | 7/12 | 10/12 | NA | 18/24 | NA | 19/21 | 30/30 | NA | Adequate |
| Scale of Solastalgia | Cáceres et al., 2022 | 30/30 | 15/27 | 10/12 | 10/12 | NA | 0/24 | 13/15 | 34/36 | 69/78 | NA | Adequate |

| | | | | | | | | | | | | |
|---|---|---|---|---|---|---|---|---|---|---|---|---|
| Validation of CCAS | Innocenti et al., 2021 | 28/30 | NA | 10/12 | 10/12 | 15/15 | 22/24 | NA | 19/21 | 28/30 | 26/36 | Good |
| Validation of CCAS | Wullenkord et al., 2021 | 30/30 | NA | 9/12 | 6/9 | 3/12 | 0/24 | NA | 18/18 | 27/27 | 26/36 | Adequate |
| Validation of CCAS | Cruz et al., 2022 | 28/30 | 5/27 | 10/12 | 10/12 | NA | 0/24 | NA | 19/21 | 28/30 | NA | Adequate |
| Validation of CCAS | Larionow, 2021 | 26/30 | NA | 10/12 | 10/12 | 10/18 | 0/24 | NA | 19/21 | 28/30 | 30/36 | Adequate |
| Validation of CCAS | Mouguiama-Daouda et al., 2022 | 30/30 | NA | 10/12 | 10/12 | 9/12 | 0/24 | NA | 19/21 | 28/30 | 31/36 | Adequate |
| Validation of CCAS | Simon et al., 2022 | 27/30 | NA | 10/12 | 10/12 | 9/18 | 0/24 | NA | 13/15 | 13/15 | NA | Adequate |
| Validation of CCAS | Jang et al., 2023 | 27/30 | NA | 10/12 | 10/12 | 6/15 | 22/24 | NA | 19/21 | 30/30 | 30/36 | Adequate |
| Validation of CCAS | Tam et al., 2023 | 30/30 | NA | 10/12 | 9/12 | 16/18 | 0/24 | NA | 21/21 | 30/30 | 14/36 | Adequate |
| Validation of CCAS | Wu et al., 2023 | 30/30 | 13/27 | 10/12 | 10/12 | NA | 0/24 | NA | 19/21 | 28/30 | NA | Adequate |
| Validation of HEAS | Uzun et al., 2022 | 27/30 | NA | 10/12 | 10/12 | 0/18 | 0/24 | NA | 0/21 | 0/30 | 27/36 | Doubtful |
| Validation of HEAS | Pavani et al., 2023 | 30/30 | NA | 8/12 | 10/12 | 0/18 | 0/24 | NA | 19/21 | 28/30 | 11/36 | Adequate |
| Validation of CCAS and HEAS | Hogg et al., 2023 | 29/30 | NA | 10/12 | 7/12 | 16/18 | 0/24 | NA | 32/36 | 50/54 | NA | Adequate |

*Note.* This table shows the results of COSMIN Study Design checklist for Patient-Reported Outcome Measurement (PROM) instruments filled out for each instrument included in the review. The first seven rows present the assessment of scale development studies. Measurement properties evaluated in the second column include the research aim, and clear descriptions of the constructs, structure of the PROM, existing evidence of the PROM, the context of use, and the target population.

* These three questionnaires were assessed simultaneously as they were tested in the same sample by following the same methodology.

| Name of the scale | Source | PROM development | Content validity | Structural validity | Internal consistency | Measurement invariance | Reliability | Measurement error | Criterion validity | Construct validity | Responsiveness | Overall Risk of Bias Rating |
|---|---|---|---|---|---|---|---|---|---|---|---|---|
| Environmental Worry Scale | Bowler & Schwarzer, 1991 | 17/105 | 0/93 | 0/9 | 3/9 | NA | 3/15 | NA | NA | 21/21 | 21/21 | High |
| Environmental Distress Scale | Higginbotham et al., 2007 | 24/105 | 0/93 | 8/9 | 9/9 | NA | 15/15 | NA | NA | 9/9 | 9/9 | Moderate |
| Climate Change Distress Scale | Searle & Gow, 2010 | 15/105 | 0/93 | 8/9 | 9/9 | NA | 3/15 | NA | NA | 12/12 | 12/12 | Moderate |
| Climate Change Anxiety Scale (CCAS) | Clayton & Karazsia, 2022 | 17/105 | 0/93 | 9/9 | 9/9 | NA | 0/15 | NA | NA | 12/12 | 12/12 | Moderate |
| Hogg Eco-Anxiety Scale (HEAS) | Hogg et al, 2021 | 21/105 | 12/93 | 7/9 | 9/9 | NA | 15/15 | NA | NA | 12/12 | 12/12 | Moderate |
| Eco-Anxiety, Eco-Guilt,and Ecological Grief Questionnaires* | Ágoston et al., 2022 | 36/105 | 17/93 | 9/9 | 9/9 | NA | 4/15 | NA | NA | 12/12 | 12/12 | Moderate |
| Climate Change Worry Scale | Stewart, 2021 | 15/105 | 0/93 | 8/9 | 9/9 | NA | 9/15 | NA | NA | 12/12 | 12/12 | Moderate |

| | | | | | | | | | | | | |
|---|---|---|---|---|---|---|---|---|---|---|---|---|
| Scale of Solastalgia | Cáceres et al., 2022 | 18/105 | 0/93 | 8/9 | 9/9 | NA | 0/15 | NA | 6/6 | 9/9 | 9/9 | Moderate |
| Validation of CCAS | Innocenti et al., 2021 | 15/15 | NA | 9/9 | 9/9 | 12/12 | 15/15 | NA | NA | 12/12 | 12/12 | Low |
| Validation of CCAS | Wullenkord et al., 2021 | 15/15 | NA | 9/9 | 9/9 | 1/12 | 3/15 | NA | NA | 12/12 | 12/12 | Moderate |
| Validation of CCAS | Cruz et al., 2022 | 14/15 | NA | 9/9 | 9/9 | NA | 3/15 | NA | NA | 12/12 | 12/12 | Low |
| Validation of CCAS | Larionow, 2021 | 13/15 | NA | 9/9 | 9/9 | 3/12 | 3/15 | NA | NA | 12/12 | 12/12 | Moderate |
| Validation of CCAS | Mouguiama-Daouda et al., 2022 | 15/15 | NA | 9/9 | 9/9 | 3/12 | 0/15 | NA | NA | 12/12 | 12/12 | Moderate |
| Validation of CCAS | Simon et al., 2022 | 12 of 15 | NA | 9/9 | 9/9 | 3/12 | 3/15 | NA | NA | 6/6 | 6/6 | Moderate |
| Validation of CCAS | Jang et al., 2023 | 12 of 15 | NA | 9/9 | 9/9 | 3/12 | 15/15 | NA | NA | 12/12 | 12/12 | Low |
| Validation of CCAS | Tam et al., 2023 | 15/15 | NA | 9/9 | 9/9 | 12/12 | 3/15 | NA | NA | 12/12 | 12/12 | Low |
| Validation of CCAS | Wu et al., 2023 | 15/15 | NA | 9/9 | 9/9 | NA | 3/15 | NA | NA | 12/12 | 12/12 | Low |
| Validation of HEAS | Uzun et al., 2022 | 12 of 15 | NA | 9/9 | 9/9 | 3/12 | 4/15 | NA | NA | 1 of 12 | 1/12 | High |
| Validation of HEAS | Pavani et al., 2023 | 15/15 | NA | 9/9 | 9/9 | 1/12 | 3/15 | NA | NA | 12/12 | 12/12 | Moderate |
| Validation of CCAS and HEAS | Hogg et al., 2023 | 14/15 | NA | 9/9 | 6 of 9 | NA | 6/15 | NA | NA | 12/12 | 12/12 | Low |

*Note*. This table shows the results of COSMIN Risk of Bias (RoB) Checklist for Patient-Reported Outcome Measurement (PROM) instruments filled out for each instrument included in the review.
